# The epidemiology of human *Taenia solium* infections: a systematic review of the distribution in Eastern and Southern Africa

**DOI:** 10.1101/2022.12.21.22283765

**Authors:** Gideon Zulu, Dominik Stelzle, Kabemba E. Mwape, Tamara M. Welte, Hilde Strømme, Chishimba Mubanga, Wilbroad Mutale, Annette Abraham, Alex Hachangu, Veronika Schmidt, Chummy. S. Sikasunge, Isaac. K. Phiri, Andrea S. Winkler

## Abstract

**Background:** *Taenia solium* is a tapeworm that causes taeniosis in humans and cysticercosis in humans and pigs. Within Eastern and Southern Africa (ESA), there are many countries in which information on the presence of human taeniosis and cysticercosis is missing. This systematic review aimed to describe the current information available and gaps in the epidemiology of human *T. solium* infections in ESA.

**Methods/Principle Findings:** Scientific literature published between 1^st^ January 2000 and 20^th^ June 2022 in international databases [MEDLINE (Ovid), Embase (Ovid), Global Health (Ovid), Scopus (Elsevier), African Index Medicus (via WHO Global Index Medicus), and Open Grey] was systematically reviewed for ESA following the PRISMA approach. The study area included 27 countries that make up the ESA region. Information on either taeniosis, cysticercosis or NCC was available for 16 of 27 countries within the region. Most case reports for cysticercosis and NCC were from South Africa, while Tanzania had the most aggregated cysticercosis reports. Eleven countries reported on NCC with seven countries reporting data on NCC and epilepsy. Unconfirmed human T. solium taeniosis cases were reported in nine countries while two countries (Madagascar and Zambia) reported confirmed T. solium cases. The cysticercosis seroprevalence ranged between 0.99 - 40.8% on antigen (Ag) tests and between 1.7 - 45.3% on antibody (Ab) tests, while NCC- suggestive lesions on brain CT scans showed a prevalence range between 1.0 - 76%. The human taeniosis prevalence based on microscopy ranged between 0.1 - 14.7%. Based on Copro Ag-ELISA studies conducted in Kenya, Rwanda, Tanzania, and Zambia, the highest prevalence of 19.7% was reported in Kenya

**Conclusions:** Despite the public health and economic impact of *T. solium* in ESA, there are still large gaps in knowledge about the occurrence of the parasite, and the resulting One Health disease complex, and monitoring of *T. solium* taeniosis and cysticercosis is mostly not in place

**Author summary:** *Taenia solium* is a tapeworm that causes three diseases, taeniosis in humans and cysticercosis in humans and pigs. Neurocysticercosis, which occurs when the central nervous system is involved has been associated with up to 57% of epilepsy cases in sub–Saharan Africa. Diagnosing neurocysticercosis among people with epilepsy is vital to prevent further morbidity and mortality from the disease as well as to reduce the negative socio-cultural beliefs associated with epilepsy. Within Eastern and Southern Africa, there are many countries in which information on the presence of human taeniosis, cysticercosis and neurocysticercosis is missing. This systematic review aimed to describe the current information available and gaps in the epidemiology of human *T. solium* infections in Eastern and Southern Africa. We found that Information on either taeniosis, cysticercosis or NCC was available only for 16 of 27 countries within the region. We also found that most of the studies on *T. solium* taeniosis, cysticercosis and neurocysticercosis within the region have been done in Kenya, Madagascar, Mozambique, Rwanda, South Africa, Tanzania and Zambia. Understanding the epidemiology of *T. solium* infections is essential for monitoring, prevention and control of the disease complex in a One Health approach.

## Introduction

### Rationale

*Taenia solium* is a tapeworm that causes taeniosis in humans and cysticercosis in humans and pigs. The life cycle of *T. solium* involves pigs as intermediate hosts (cysticercosis) while humans are definitive hosts (taeniosis). Humans may also act as accidental intermediate hosts when larvae of the parasite settle in muscles, subcutaneous or organ tissues causing human cysticercosis (HC). If they lodge in the central nervous system (CNS), including the brain and the spinal cord, the disease is called neurocysticercosis (NCC) [1, 2]. An individual may also have cysticerci in the CNS as well as in other parts of the body, which is referred to as (neuro) cysticercosis. NCC may be asymptomatic, but it can also cause various neurological signs/symptoms such as epileptic seizures and epilepsy, chronic progressive headache, focal neurological deficits, signs and symptoms of increased intracranial pressure and in rare cases, death [3–5]. Globally, NCC is estimated to be responsible for 30% of cases of acquired epilepsy in endemic areas [6] and 22% in sub-Saharan Africa [7]. However, this proportion depends on the infection pressure of *T. solium* and thus may vary considerably.

*T. solium* is common in Asia and Latin America, especially in areas where pigs are reared under free-ranging conditions, where pork is eaten and where hygiene is limited [8, 9]. Hence, *T. solium* is also likely endemic in most African countries [10]. Nevertheless, information on the endemicity of *T. solium* is limited and there are many countries from which no published information is available for either human or porcine cysticercosis. The purpose of this review was to gather all published information on human *T. solium* infections (*T. solium* taeniosis/(neuro)cysticercosis (TSTC)) in eastern and southern Africa (ESA) within the period 1st January 2000 and 20th June 2022 and describe the gaps in the epidemiology of TSTC in this area.

## Methods

The systematic review was conducted following the Preferred Reporting Items for Systematic Reviews and Meta-Analyses (PRISMA) guidelines [11] (S1 Table). The review was registered with the International Prospective Register of Systematic Reviews (PROSPERO) (registration number: CRD 42022343072).

### Search strategy

All published articles were searched using the electronic databases, MEDLINE (Ovid), Embase (Ovid), Global Health (Ovid), Scopus (Elsevier), African Index Medicus (via WHO Global Index Medicus), and Open Grey. The reference lists of included studies were also scanned to identify further eligible documents. The searches were conducted on 20^th^ June 2022.

For all questions subject headings (where applicable) and text words describing *T. solium* taeniosis/cysticercosis/neurocysticercosis and prevalence, epidemiology, control, elimination, or eradication were searched. The search strategy was intended to obtain all available literature on human taeniosis, cysticercosis and neurocysticercosis in ESA conducted and published in the last 22 years. Grey literature was also searched for any relevant publications. The search terms for all databases can be found in (S1 File).

### Procedures

#### Data management

Literature search results were uploaded to the bibliographic software Mendeley Desktop and Covidence. Duplicates were removed based on author names and titles. Screening questions based on the inclusion and exclusion criteria were developed and citation abstracts and full-text articles were uploaded.

#### Selection process

The selection process was conducted in two stages. Firstly, titles and abstracts were screened independently by two reviewers (GZ and CM) and then full-text reports were obtained by the same reviewers for a detailed assessment with respect to the inclusion criteria. For records that referred to the same study, only one was selected.

#### Data collection process

The data were collected using standardized data collection forms in Microsoft Excel (S2 File). Data extracted included demographic information, methodology, and all the relevant reported outcomes. GZ carried out the data extraction with verification by CM.

#### Data items

Predefined tables summarizing individual cases included the year of diagnosis, age, gender, the country where cases were detected, diagnostic method, and location of the lesion (for cysticercosis). Tables summarizing aggregated cases or prevalence data included country, level of data collection (e.g. national or regional), timeframe, number of cases (or prevalence proportion), number of people tested, and diagnostic method.

### Eligibility criteria

#### Inclusion criteria

We included all studies describing human TSTC in ESA, which was defined as the area covered by the following countries/territories: Angola, Botswana, Burundi, Comoros, Djibouti, Eritrea, Ethiopia, Kenya, Lesotho, Madagascar, Malawi, Mauritius, Mayotte, Mozambique, Namibia, Reunion, Rwanda, Seychelles, Socotra, Somalia, Somaliland, South Africa, Eswatini (former Kingdom of Swaziland), Tanzania, Uganda, Zambia and Zimbabwe. Included in this review were studies that describe the epidemiology, disease occurrence, burden, prevalence, incidence, prevention and control of TSTC in any of the twenty-seven countries of ESA. All observational studies including case series and case reports and systematic reviews of such studies were included.

Only studies conducted between 1^st^ January 2000 and 20^th^ June 2022 were considered. No language limits were imposed on the search, although studies in languages other than English were only included if they could be adequately translated by using Google Translate.

#### Exclusion criteria

We excluded studies that did not concern *T. solium,* did not concern humans, did not report data from within ESA and studies conducted before 1^st^ January 2000.

### Risk of bias assessment

The risk of bias in the studies was assessed using items recommended by the Agency for Health Care Research and Quality (AHRQ) [12]. This assessment covered baseline characteristics, inclusion-exclusion criteria, confounding and modifying variables, performance bias, attrition bias, detection bias and reporting bias. Each of these factors was rated and categorized as low, or high risk of bias. If the information available was insufficient, the risk of bias was considered ’unsure’.

### Statistical analyses

No statistical pooling of results was conducted and the findings of the review are presented in a narrative synthesis with tables and figures to aid data presentation. Prevalence data if not already provided were calculated using the numerator and denominator of the study sample. Data were analyzed separately for taeniosis, cysticercosis, and neurocysticercosis. For the latter two, data were also presented separately for the general population and people with epilepsy.

### Ethical considerations

Ethical recommendation was not required for this review.

## Results

### Study selection

Our search yielded 2114 records of which 1198 were duplicates. Through screening of titles and abstracts, 776 (85%) records were excluded and for a further 8, no full text was found. Full-text assessment with respect to the inclusion criteria was performed on 132 records of which 19 (14%) were excluded (17 reported data described in other included studies, one did not concern *T. solium*, and one reported results outside the scope of the study). One hundred and thirteen (113) studies were finally included in the review. Of those, 49 were concerned with only (neuro) cysticercosis, 57 with taeniosis, and 7 with both cysticercosis and taeniosis. Of the 113 reports reviewed, 25 were case reports, 82 were cross-sectional, 3 cohort and 3 case-control studies. The flow diagram for the search is shown in (Fig 1). The reference for included studies is shown in (S2 Table).

**Fig 1.**
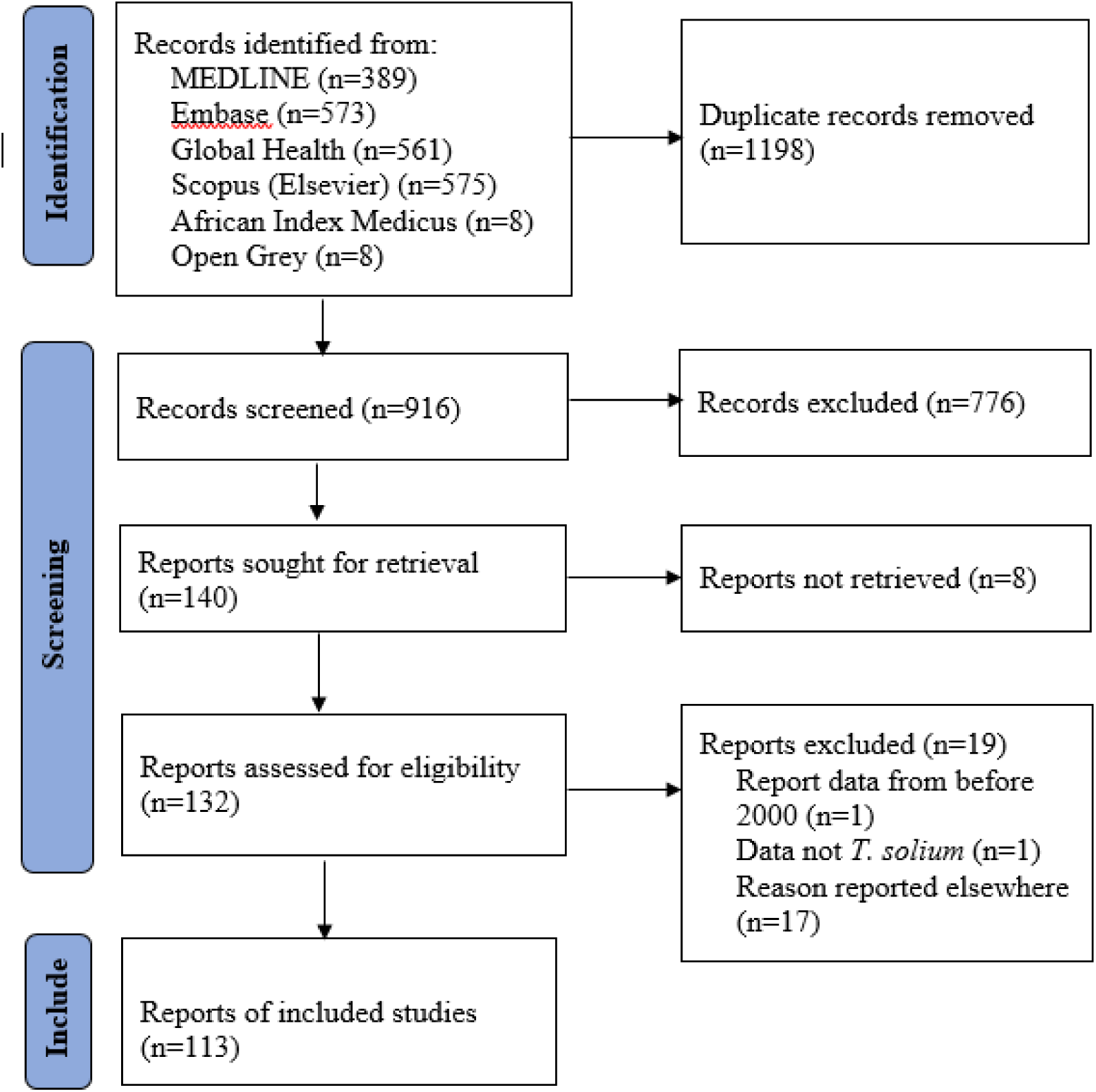
PRISMA flow diagram

### Risk of bias

The risk of bias assessment revealed that most studies had a low risk of reporting, detection, attrition and performance bias. However, 7 studies had a high risk of performance bias, 20 had a high risk of bias due to inadequate inclusion and exclusion criteria, and 45 articles had a high risk of bias due to confounding and modifying variables.

### Results of individual studies

#### Human (neuro) cysticercosis

Information on human (neuro) cysticercosis was obtained from 62 sources. In total, 25 (40%) records provided individual information on NCC and CC. Twelve (48%) of these records were on NCC only, seven (28%) on CC and six (24%) on both NCC and CC. Thirty-seven (60%) records reported aggregated information on NCC and CC. Ten (27%) of these records were on NCC only, twenty-one (57%) on CC only, and six (16%) on both NCC and CC. Information on human neuro(cysticercosis) was available for 14 of 27 ESA countries. No reports were available for Angola, Botswana, Comoros, Djibouti, Eritrea, Ethiopia, Lesotho, Mayotte, Reunion, Seychelles, Socotra, Somaliland and Somalia.

#### Human cysticercosis case reports

Individual human CC cases during the period 2000 to 2022 were reported in 13 hospital case records from six of the ESA countries with, South Africa reporting four cases; Madagascar, Malawi, Rwanda and Tanzania reporting two cases each and Mauritius reporting one case (Table 1). Individual NCC cases were reported in 18 hospital records from eight of the ESA countries with South Africa reporting eight cases while Madagascar, Malawi and Zambia reported two cases each. Mozambique, Eswatini, Tanzania and Zimbabwe reported one case each (Table 2). All reports were based on individual patients presenting with various signs/symptoms, and investigations leading to their diagnosis of either CC or NCC. Twelve (48%) of the 25 case reports described only NCC diagnosed through the use of brain computed tomography (CT) scan (7) and magnetic resonance imaging (MRI) (5), while seven (28%) presented with CC affecting other organs i.e. bronchus, breast, abdomen, neck and subcutaneous tissue diagnosed through histopathology and other investigations. Six (24%) case reports were on patients who presented with both NCC and CC affecting other organs (disseminated CC). At a district hospital in Rwanda ten cases of cutaneous CC were reported with the youngest being only 2 months old [13]. The oldest reported case of NCC was from South Africa in an 86-year-old female with a pharyngeal cyst, cutaneous cysts and active brain cysts [14]. The median age in years was 41 (17 -48 year). Twenty-three (92%) of the case reports had information on the affected gender with 54% of the reported cases being female and 46% being male.

**Table 1.** Individual human cysticercosis case reports as detected through histopathology of sample specimens from patients in eastern and southern Africa (2000 - 2022)

| Country | Age (yrs.) | Gender | Sample | Location of pathogen | Comment | Reference |
| --- | --- | --- | --- | --- | --- | --- |
| Madagascar | 20 | Male | Bronchial specimen | Bronchus | A 20-year-old male who presented with hemoptysis following bronchial cysticercosis. | [50] |
|  | 15 | Female | Breast lump | Breast | Breast cysticercosis in a 15-year-old girl. | [51] |
| Malawi | 41 | Female | Skin nodule | Subcutaneous all over the body* | A 41-year-old patient on ARVs with disseminated cysticercosis and more than 50 cysts in the brain. | [52] |
|  | 56 | Female | Biopsy of neck mass | Left anterior and posterior triangles of the neck | A 56-year-old woman with a history of gradual but progressive growing left sided neck swelling for the past 25 years. | [53] |
| Mauritius | 22 | Female | Excision biopsy | Upper part of right breast | A 22-year-old woman with history of painless mobile swelling in the right side of the breast. | [54] |
| Rwanda | 2 months - 65 yrs. | - | Anatomical department files | Subcutaneous nodules | A case report of 10 cases of cutaneous cysticercosis confirmed by histopathology in the anatomical pathology department at a district hospital (Nyamasheke) in eastern Rwanda. | [13] |
|  | 46 | Female | Subcutaneous nodule | Subcutaneous nodules on trunk and extremities | A 46-year-old woman who presented with a two-week history of bilateral lower limb weakness, causing difficulty walking. | [55] |
| South Africa | 42 | Male | Biopsy of subcutaneous nodule | Extensive calcifications in the muscle and subcutaneous tissue. Active cysts within myocardium<sup>#</sup> | A 42-year-old man with a rare case of myocardial cysticercosis with multiple calcifications in the muscle and subcutaneous tissues. | [56] |
|  | 86 | Female | Excised cyst from pharynx<br>Subcutaneous nodule biopsy | Left posterior oropharynx<br>Subcutaneous nodules on the back | An 86-year-old woman with cysts in the left posterior oropharynx, brain and subcutaneous tissues on the neck and back. | [14] |

| Country | Age<br>(yrs.) | Gender | Sample | Location of pathogen | Comment | Reference |
| --- | --- | --- | --- | --- | --- | --- |
|  | 49 | Male | Excisional<br>nodule biopsy | Subcutaneous nodules<br>on the trunk and upper<br>extremities | A 49-year-old man with generalized cysticercosis. | [57] |
|  | 8 | Nr | Cyst from<br>pleural<br>effusion | Right lung | An 8-year-old child with pulmonary cysticercosis who<br>presented with a large right pleural effusion. | [58] |
| Tanzania | 4 | Male | Subcutaneous<br>sample biopsy | Sub mandibular and<br>cervical region and<br>abdomen | A 4-year-old HIV positive child with disseminated<br>cysticercosis. | [59] |
|  | 48 | Male | Subcutaneous<br>nodule | Neck | A 48-year-old male driver with disseminated cysticercosis<br>treated as malaria. | [60] |

ARV, Antiretroviral medication; HIV, Human immunodeficiency virus; Yrs., Years.
\*detected through musculoskeletal ultrasound of nodules
#detected through chest and soft tissue Rontgen radiation (X-rays)

**Table 2.** Individual human neurocysticercosis (NCC) case reports as detected by Brain CT in eastern and southern Africa (2000 - 2022)

| Country | Age<br>(yrs.) | Gender | Location of pathogen | Comment | Reference |
| --- | --- | --- | --- | --- | --- |
| Eswatini | 40 | Male | Multiple intra-parenchymal cysts, mural scoleces, calcified granulomas, right sided and parafalcine septated subdural collections, pathognomonic of racemose, stage I and stage IV NCC | A 40-year-old male with a history of chronic headache, unsteady gait and progressive deterioration in vision. | [69] |
| Madagascar | 71 | Male | Not described* | A 71-year-old man with dysarthria and memory problems. | [61] |
|  | 31 | Male | Two ring enhancing lesions in the left capsulo-lenticular.<sup>†</sup> | A 31-year-old man presenting with dysarthria, blurred vision and monoparesis of the right upper limb 7 weeks after starting anti-tuberculosis medication. | [62] |
| Malawi | 58 | Female | Multicystic lesion in the left temporal lobe<sup>¶#</sup> | A 58-year-old woman with coincidental racemose NCC who was being treated for HIV associated cryptococcal meningitis, clinically and microbiologically responding to antifungal therapy. | [63] |
|  | 41 | Female | Multiple intra-parenchymal cyst in the brain | A 41-year-old patient on ARVs with disseminated cysticercosis and more than 50 cysts in brain. | [52] |
| Mozambique | 18 | Female | Multiple lesions with different stages ranging from cerebral ring-enhancing cysts with scolices to transitional or degenerative colloidal cysts to multiple parenchymal calcifications | An 18-year-old female admitted to Hospital Central de Quelimane, Zambezia province in Mozambique, 8 days after delivery with a misdiagnosis of eclampsia that turned out to be NCC. | [64] |
| South Africa | 52 | Female | Granuloma within bilateral frontal and temporal lobes; Multicystic racemose lesion in the right cerebellar hemisphere; active cyst within the left cerebellar hemisphere; hydrocephalus with blocked ventriculo- peritoneal shunt; Intraventricular cyst | A 52-year-old woman with multiple types of NCC pathology. | [65] |
|  | 42 | Male | Multiple active and calcified lesions in the brain. | A 42-year-old man with NCC and disseminated cysticercosis with myocardial involvement and with multiple calcifications in the muscle and subcutaneous regions. | [56] |
|  | 86 | Female | Active brain cysts (not specific) | An 86-year-old woman with NCC and cysts in the left posterior oropharynx and subcutaneous tissues on the neck and back. | [14] |
|  | 49 | Male | Bilateral multiple parenchymal non-enhancing cerebral cystic lesions | A 49-year-old man with generalized cysticercosis. | [57] |
|  | 46 | Female | Spinal and vertebra region. A spinal lesion at T12, causing vertebral collapse<sup>#§†</sup> | A 46-year-old HIV+ patient with NCC affecting the spine with severe pain with spasms and weakness in lower limbs. | [66] |
|  | 30 | Female | Cyst in the occipital horn of the right lateral ventricle | A 30-year-old woman with acute unilateral hydrocephalus. | [67] |
|  | 8 | - | A non-specific solitary sub centimeter calcified granuloma abutting the tentorium cerebellum | Letter to the editor of a case report on an 8-year-old child with pulmonary cysticercosis who presented to Red Cross children's hospital, Cape Town, with a large right pleural effusion. | [58] |
|  | 32 | Male | Brain with isolated cystic lesions with scolex and ring enhancing lesion in colloidal stage | A 32-year-old known PWE admitted with COVID-19 and developed recurrent epileptic seizures due to dying cysts after beginning of NCC treatment. | [68] |
|  | 42 | Female | Not described | A 42-year-old known PWE admitted with COVID-19 and NCC with poor response to treatment and in status epilepticus for a long time. |  |
| Tanzania | 48 | Male | Multiple cystic lesions with dot sign diffusely distributed in the brain, neck, chest, including the heart, abdomen, and pelvis<sup>†</sup> | A 48-year-old driver with disseminated cysticercosis treated as malaria. | [60] |
| Zambia | 48 | Female | Numerous round points with T1 hypo intense, T2 hyper intense dot-form cores seen in all cerebral slices. <sup>#</sup> | A 48-year-old woman who presented with subcutaneous nodules on the face, recurrent headache and epileptic seizures in Chambishi Zambia. | [70] |
|  | 60 | Male | Thoracic intramedullary cystic lesions. Also cystic lesions in subcutaneous tissue<sup>#†</sup> | A 60-year-old man with progressive lower extremity weakness and numbness at University Teaching Hospital in Zambia. | [71] |
| Zimbabwe | 35 | Male | A Suprasellar cystic lesion compressing the optic nerve<sup>#§</sup> | A 35-year-old man presenting with pain accompanied by mild vision loss in the right eye due to a cystic mass compressing the optic nerve. | [72] |

Ag-ELISA, Antigen based enzyme linked immunosorbent assay; ARVs, Antiretroviral medication; CSF, Cerebrospinal fluid; CT, Computed tomography; EITB,
Enzyme linked electroimmunotransfer blot; ELISA, Enzyme linked immunosorbent assay; HIV, Human immunodeficiency virus; MRI, Magnetic resonance
imaging; NCC, Neurocysticercosis; Yrs., Years.
\* Serology with Ag-ELISA and EITB also used for diagnosis
¶ CSF ELISA also used for diagnosis

#### Aggregated human cysticercosis

The number of aggregated human CC reports (community-based, cross-sectional studies) for the period 2000 to 2022 in ESA was 27 (Table 3). These were reports from 9 nine countries, the majority of which were conducted in Tanzania (n=7) followed by Madagascar (n=6) and Zambia (n=4). South Africa, Mozambique, Rwanda and Kenya had two reports each. For Burundi and Uganda one report each was available. There was great variation in the aggregated number of human CC cases across countries with over 2307 cases reported from Madagascar only. The number of human CC cases identified in other ESA countries in documents published between 2000 and 2022 is shown in (Fig 2) below.

**Fig 2.**
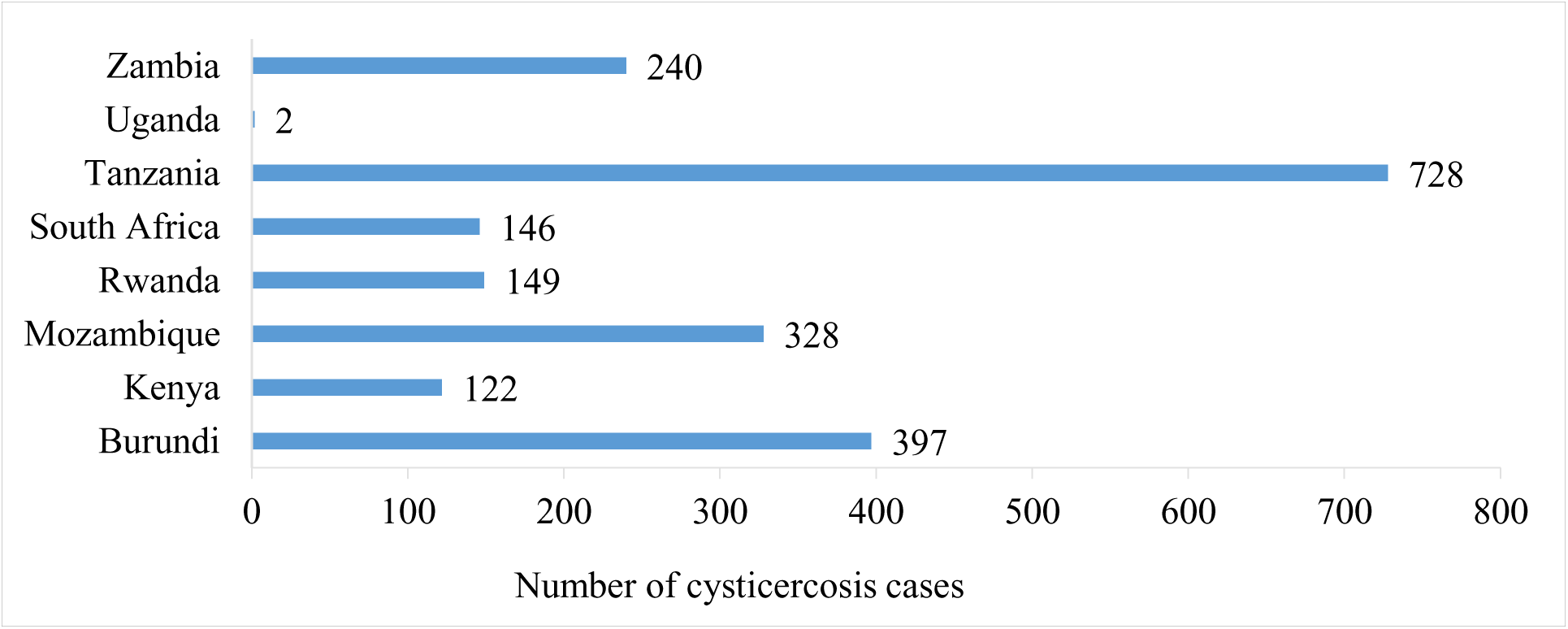
Aggregated human cysticercosis cases reported in community-based cross-sectional studies in eastern and southern Africa between the years 2000 and 2022.

**Table 3.**
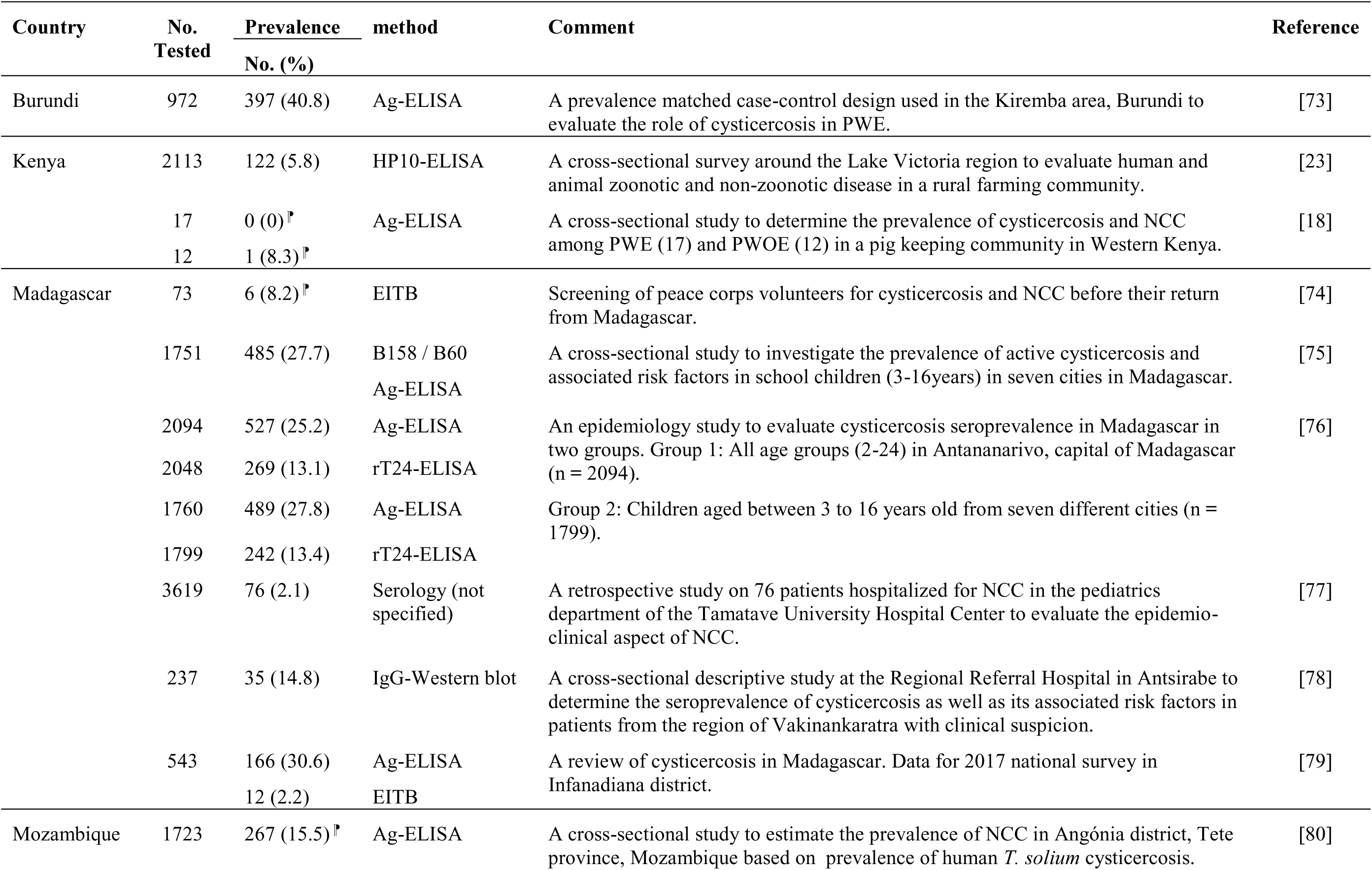
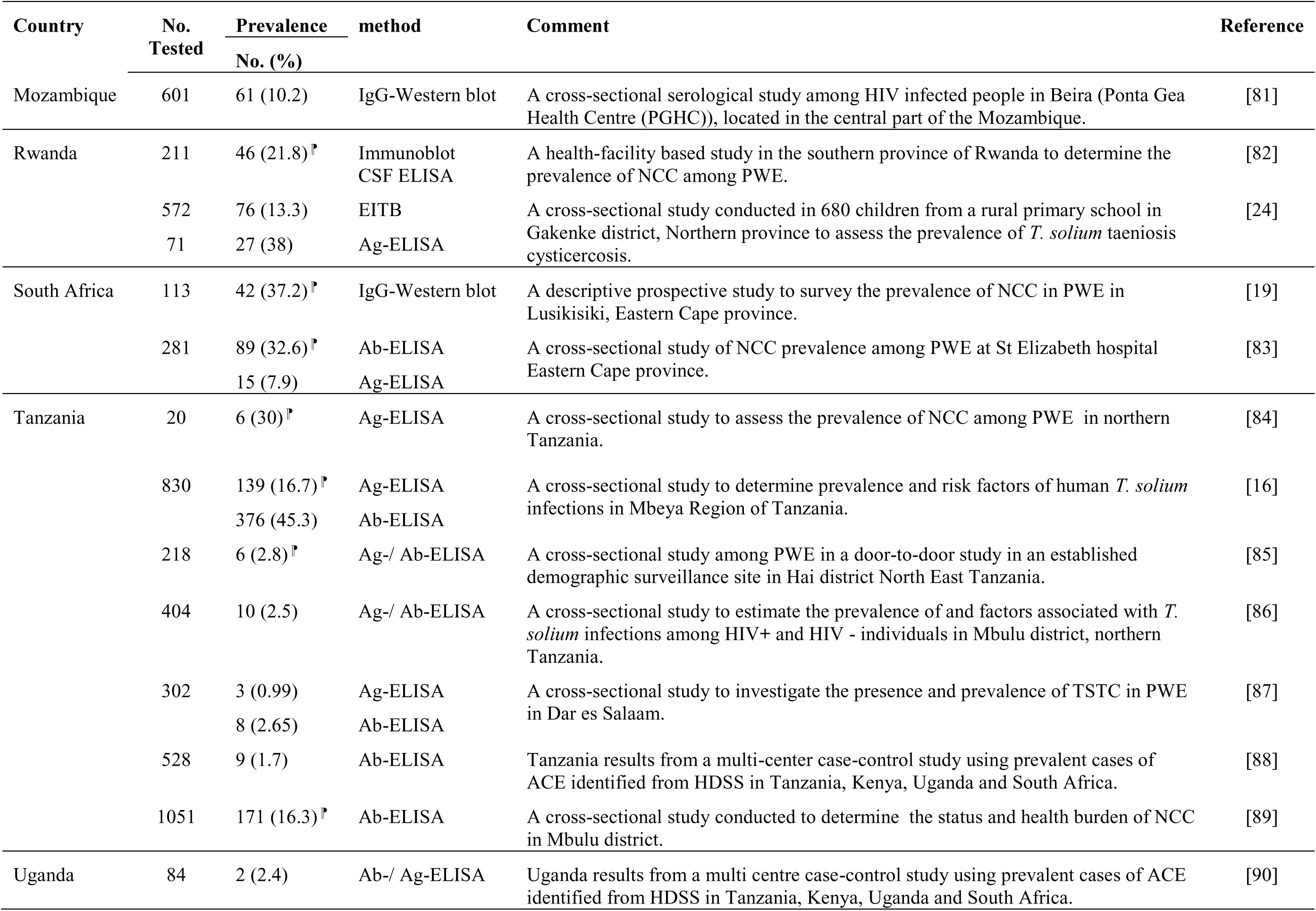
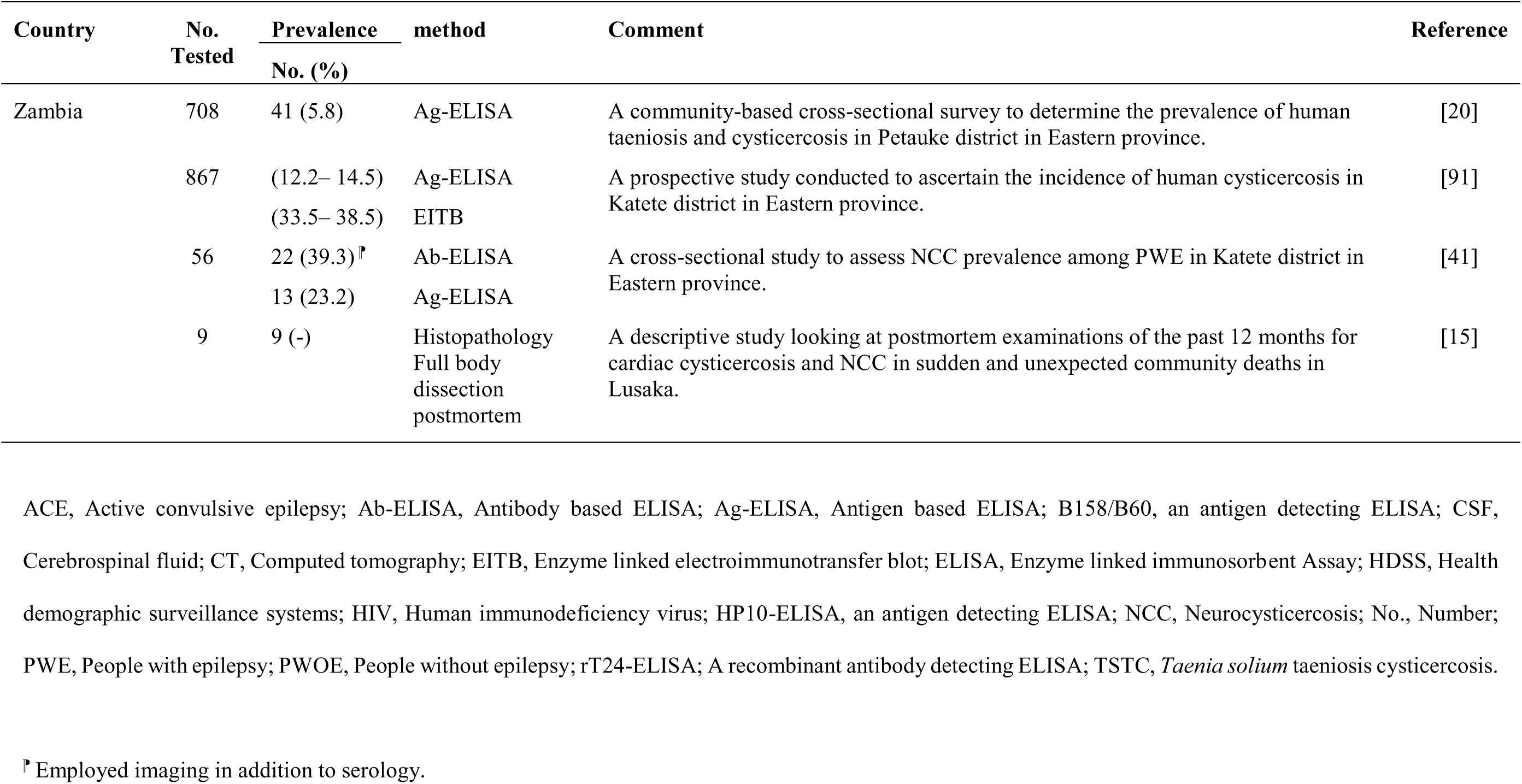
Aggregated human cysticercosis cases identified in eastern and southern Africa (2000 - 2022).

To ascertain cases of aggregated human CC, various diagnostic methods were utilized in the included reports with eight (30%) studies employing a combination of antibody and antigen (Ab/Ag) based enzyme linked immunosorbent assay (ELISA), five (19%) employed Ag-ELISA, while three reports (11%) utilized a combination of Ag-ELISA and electroimmunotransfer blot assay (EITB). One report (4%) was based on postmortem findings of CC through full-body dissection [15]. The other methods used for CC diagnosis are shown in (Fig 3).

**Fig 3.**
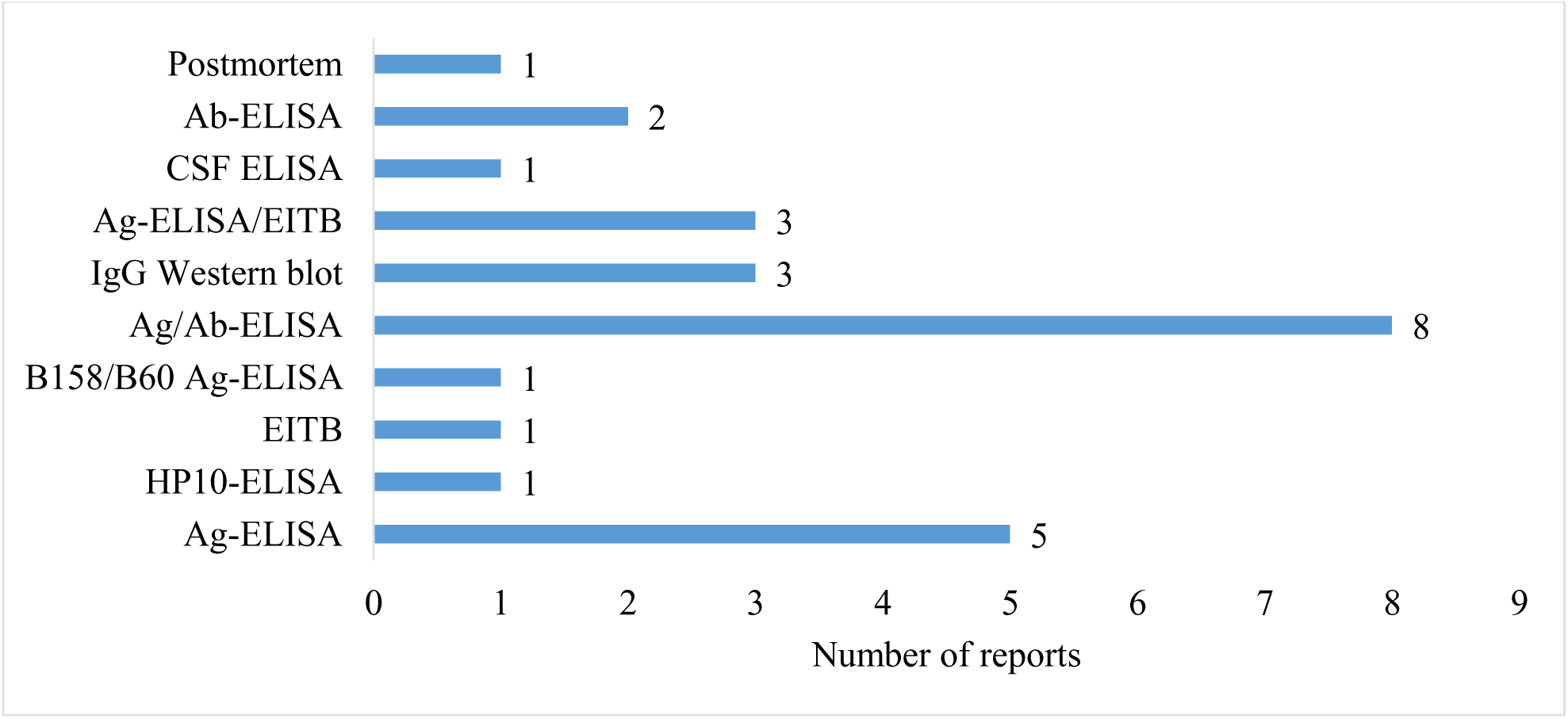
Diagnostic methods used for aggregated human cysticercosis cases in the included studies for eastern and southern Africa

The prevalence of human CC in ESA in different study populations showed wide variation within and between countries (Fig 4 and Table 3). The CC seroprevalence ranged between 0.99 – 40.8% on Ag tests and between 1.7 – 45.3% on Ab tests. The highest point prevalence was reported in Tanzania (45.3%) from a study conducted in the Mbozi area of Mbeya District [16].

**Fig 4.**
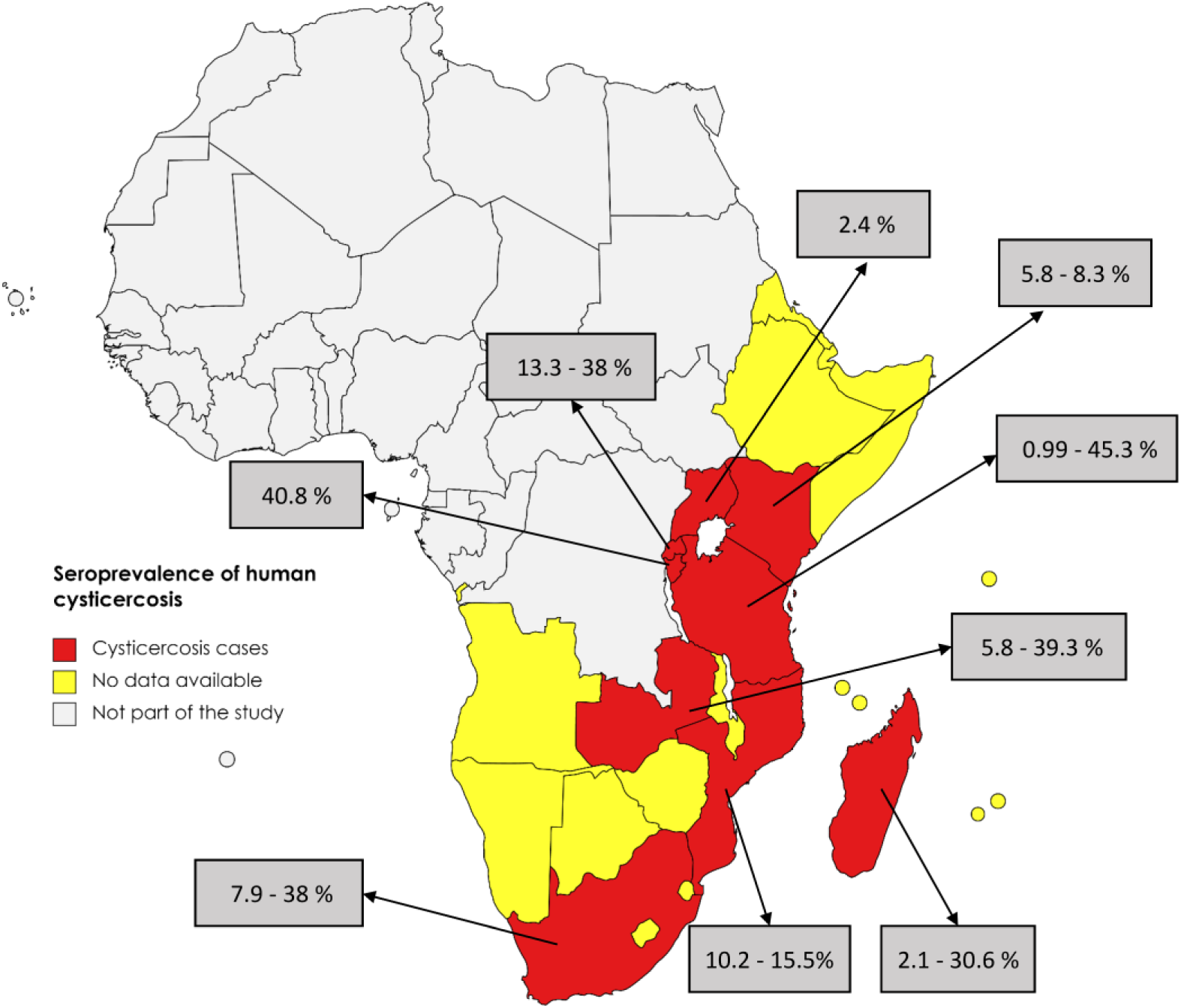
The distribution of human cysticercosis seroprevalence in the included studies for eastern and southern Africa.

For the studies conducted within Tanzania, this was an outlier value. High prevalence variation was also observed within countries as seen in Madagascar, Rwanda, South Africa and Zambia (Fig 5). The lowest point prevalence (0.99%) was also reported from Tanzania. For details on sources refer to Table 3.

**Fig 5.**
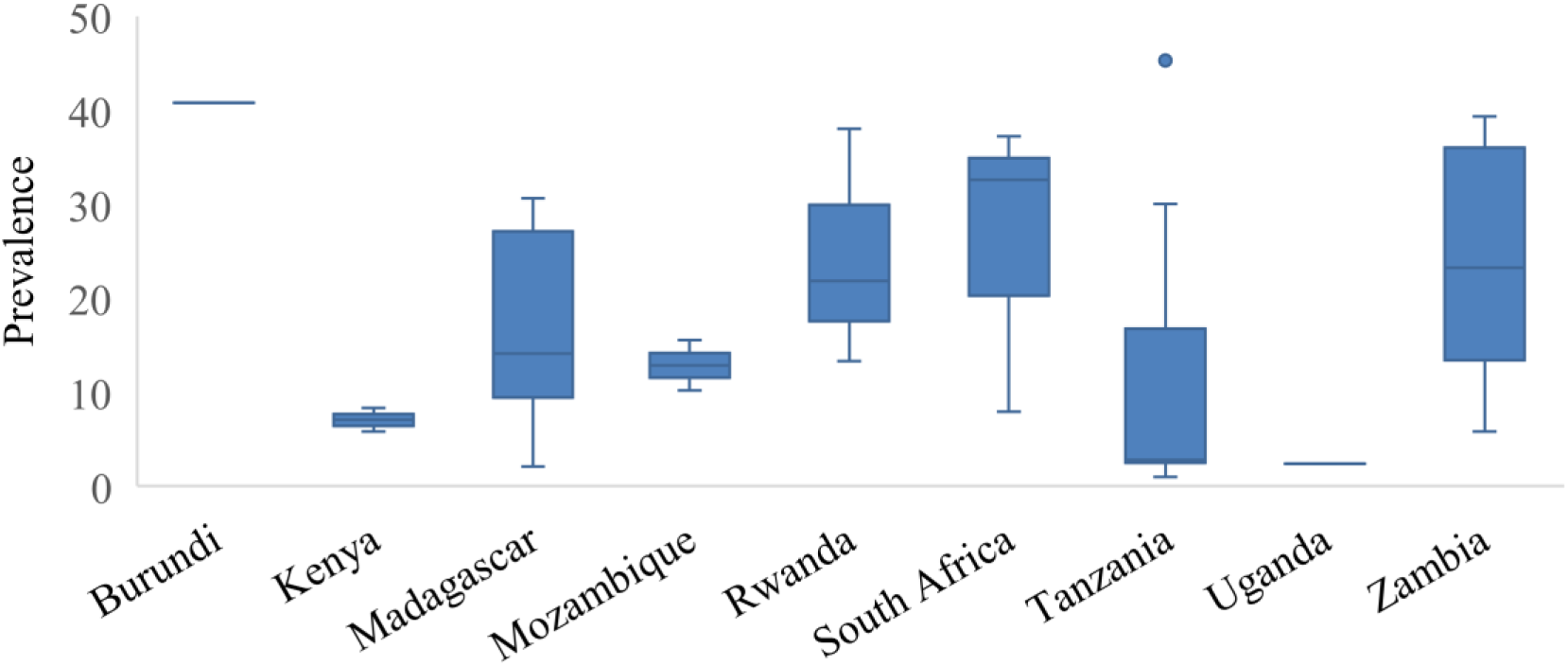
Prevalence of human cysticercosis in eastern and southern Africa based on serology data in reviewed studies with high variation observed in Rwanda, South Africa, Tanzania and Zambia

### Aggregated human neurocysticercosis

For reports presenting aggregated NCC cases (Table 4), 16 records were available. Four (25%) employed imaging (brain CT or MRI) only, whereas 11 (69%) incorporated both imaging and serological tests, and one (6%) was based on postmortem full-body dissection. The NCC- suggestive lesions on brain CT scans showed a prevalence range between 1.0 – 76% in different study populations in ESA (Fig 6 and Table 4).

**Fig 6.**
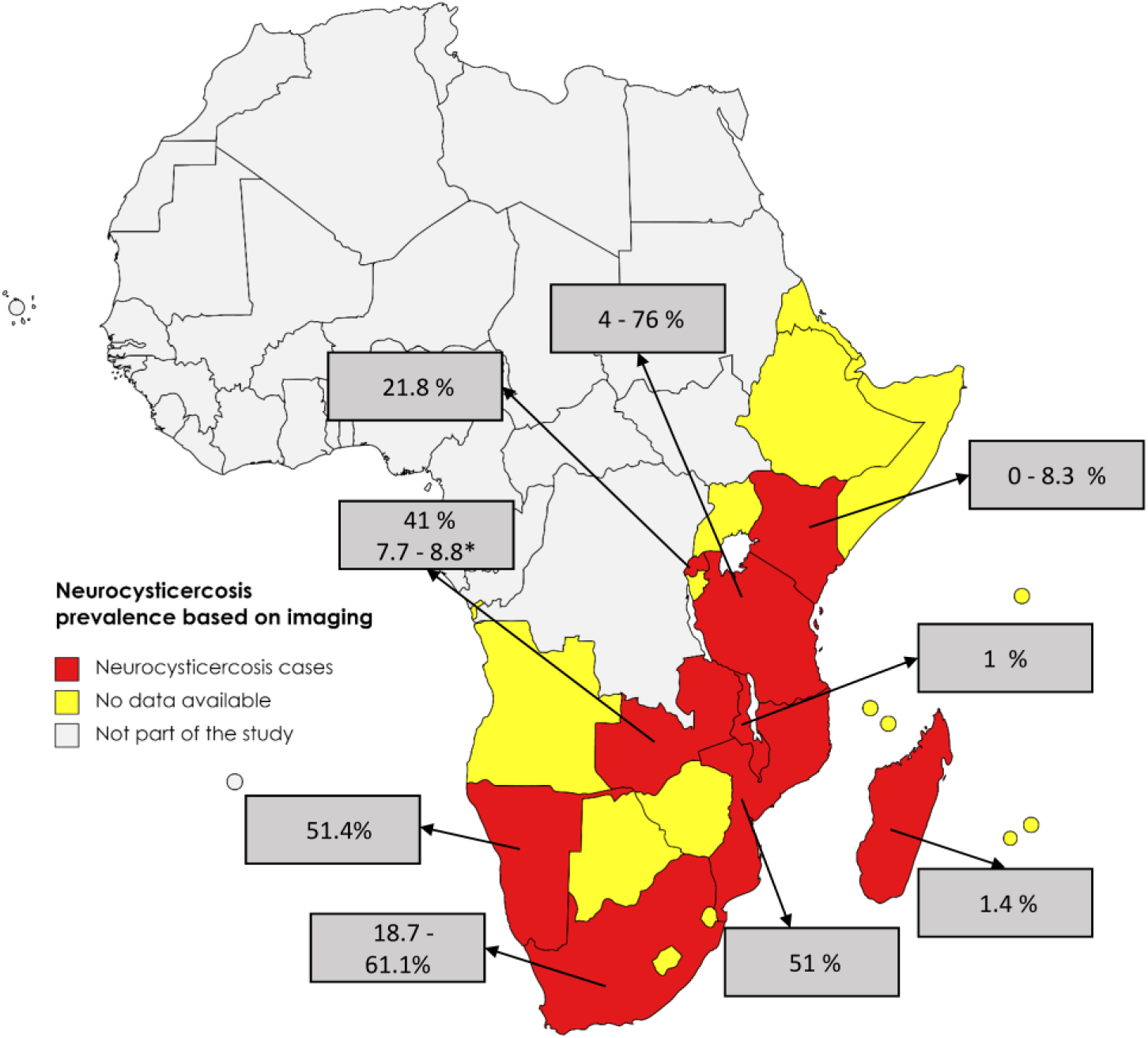
The distribution of human neurocysticercosis prevalence based on imaging data in reviewed studies in eastern and southern Africa. *Prevalence based on MRI

**Table 4.**
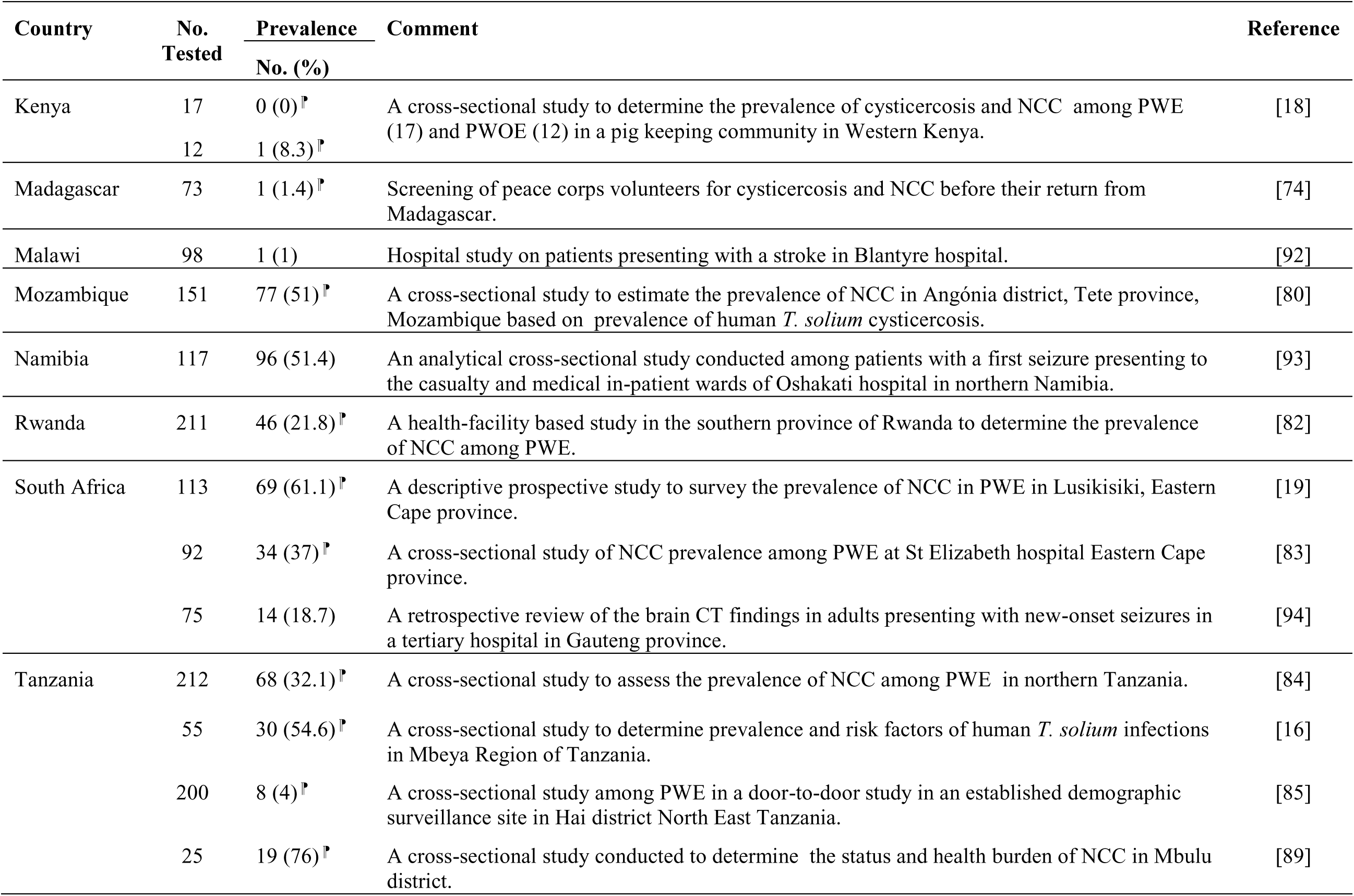
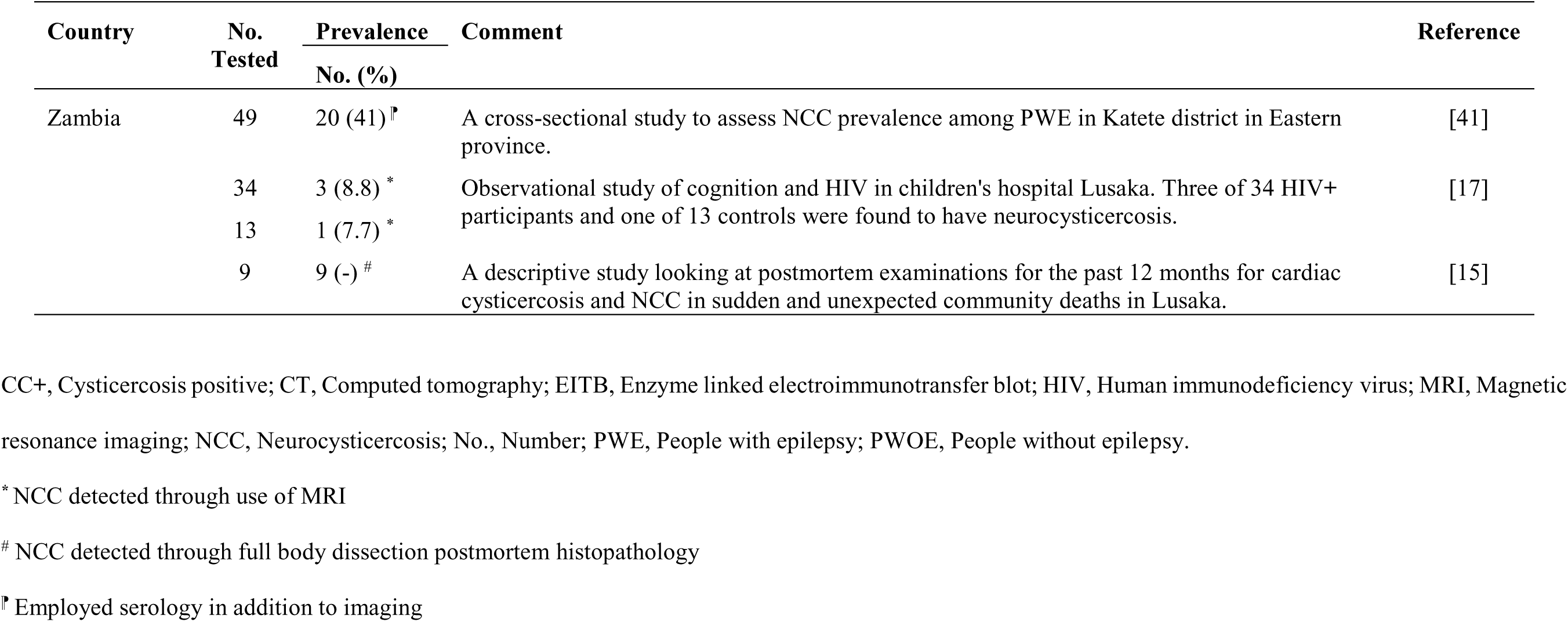
Aggregated human neurocysticercosis cases as detected by Brain CT scan in eastern and southern Africa (2000 - 2022)

One report utilized MRI to identify lesions suggestive of NCC in HIV+ children. In this report, four children (8.5% of the total) were found to have NCC, with imaging suggesting the vesicular (active) stage of NCC in all of them [17].

### Neurocysticercosis and epilepsy

Among the 16 records reporting aggregated NCC, nine (56%) reported data linking NCC and epilepsy (Table 5). Three of these records were reported from South Africa whereas one record each was reported from Kenya, Mozambique, Namibia, Rwanda, Tanzania and Zambia. Three (33%) of these NCC epilepsy studies were community-based studies and six (67%) were conducted in hospital/health facility settings. The prevalence of NCC among people with epilepsy ranged between 0 – 61%. One study in Kenya in a *T. solium* endemic area found none of the participants with epilepsy had serological evidence of cysticercosis and none had radiographic findings consistent with NCC [18]. Whereas at St Elizabeth’s Hospital, Lusikisiki, Eastern Cape, 61% of the patients presenting with epilepsy had NCC associated epilepsy, the prevalence being highest in the 10 - 19-year-old age group [19]. For details on sources refer to Table 5.

**Table 5.** Human neurocysticercosis with epilepsy as detected by Brain CT scan in eastern and southern Africa (2000 - 2022).

| Country | No<br>Tested | Prevalence<br>No (%) | Comment | Reference |
| --- | --- | --- | --- | --- |
| Kenya | 17 | 0 (0)<sup>¶</sup> | A cross-sectional study to determine the prevalence of cysticercosis and NCC among PWE in a pig keeping community in Western Kenya | [18] |
| Mozambique | 151 | 77 (51)<sup>¶</sup> | A cross-sectional study to estimate the prevalence of NCC in Angónia district, Tete province, Mozambique based on prevalence of human *T. solium* cysticercosis. | [80] |
| Namibia | 117 | 96 (51.4) | An analytical cross-sectional study conducted among patients with a first seizure presenting to the casualty and medical in-patient wards of Oshakati hospital in northern Namibia. | [93] |
| Rwanda | 211 | 46 (21.8)<sup>¶</sup> | A health-facility based study in the southern province of Rwanda to determine the prevalence of NCC among PWE. | [82] |
| South Africa | 113 | 69 (61.1)<sup>¶</sup> | A descriptive prospective study to survey the prevalence of NCC in PWE in Lusikisiki, Eastern Cape province. | [19] |
|  | 92 | 34 (37)<sup>¶</sup> | A cross-sectional study of NCC prevalence among PWE at St Elizabeth hospital Eastern Cape province. | [83] |
|  | 75 | 14 (18.7) | A retrospective review of the brain CT findings in adults presenting with new-onset seizures in a tertiary hospital in Gauteng province. | [94] |
| Tanzania | 212 | 68 (32.1)<sup>¶</sup> | A cross-sectional study to assess the prevalence of NCC among PWE in northern Tanzania. | [84] |
| Zambia | 49 | 20 (41)<sup>¶</sup> | A cross-sectional study to assess NCC prevalence among PWE in Katete district in Eastern province. | [41] |

CT, Computed tomography; NCC, Neurocysticercosis; PWE, People with epilepsy.
<sup>¶</sup> Employed serology in addition to imaging

#### Human taeniosis

A total of 64 records were identified providing information on taeniosis cases in ESA for the period 2000 to 2022 (Table 6). These records were reported in 11 out of 27 ESA countries (Fig 7) with Ethiopia reporting the most records (n=38) followed by Tanzania (n=6) and Zambia (n=5). Kenya and Uganda had three records each while Angola, Mozambique, and South Africa had two records each. Malawi, Madagascar and Rwanda had one record each. There were no cases reported from Botswana, Burundi, Comoros, Djibouti, Eritrea, Lesotho, Mauritius, Mayotte, Namibia, Reunion, Seychelles, Socotra, Somalia, Somaliland, Swaziland, and Zimbabwe.

**Fig 7.**
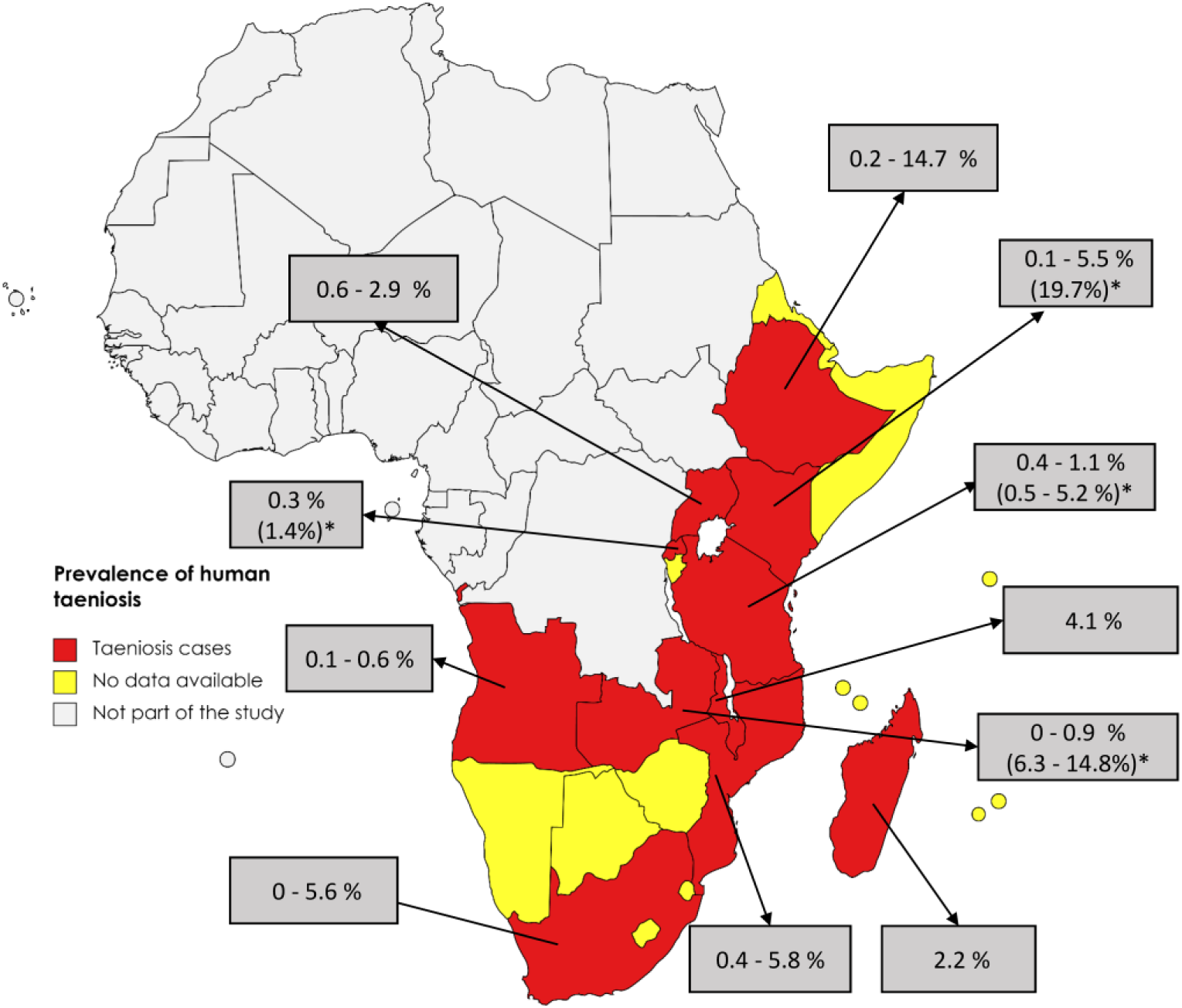
The distribution of human taeniosis in eastern and southern Africa showing prevalence data on microscopy and *prevalence based on serology (rES33-immunoblot, Ag-ELISA), and Copro-Ag ELISA)

**Table 6.**
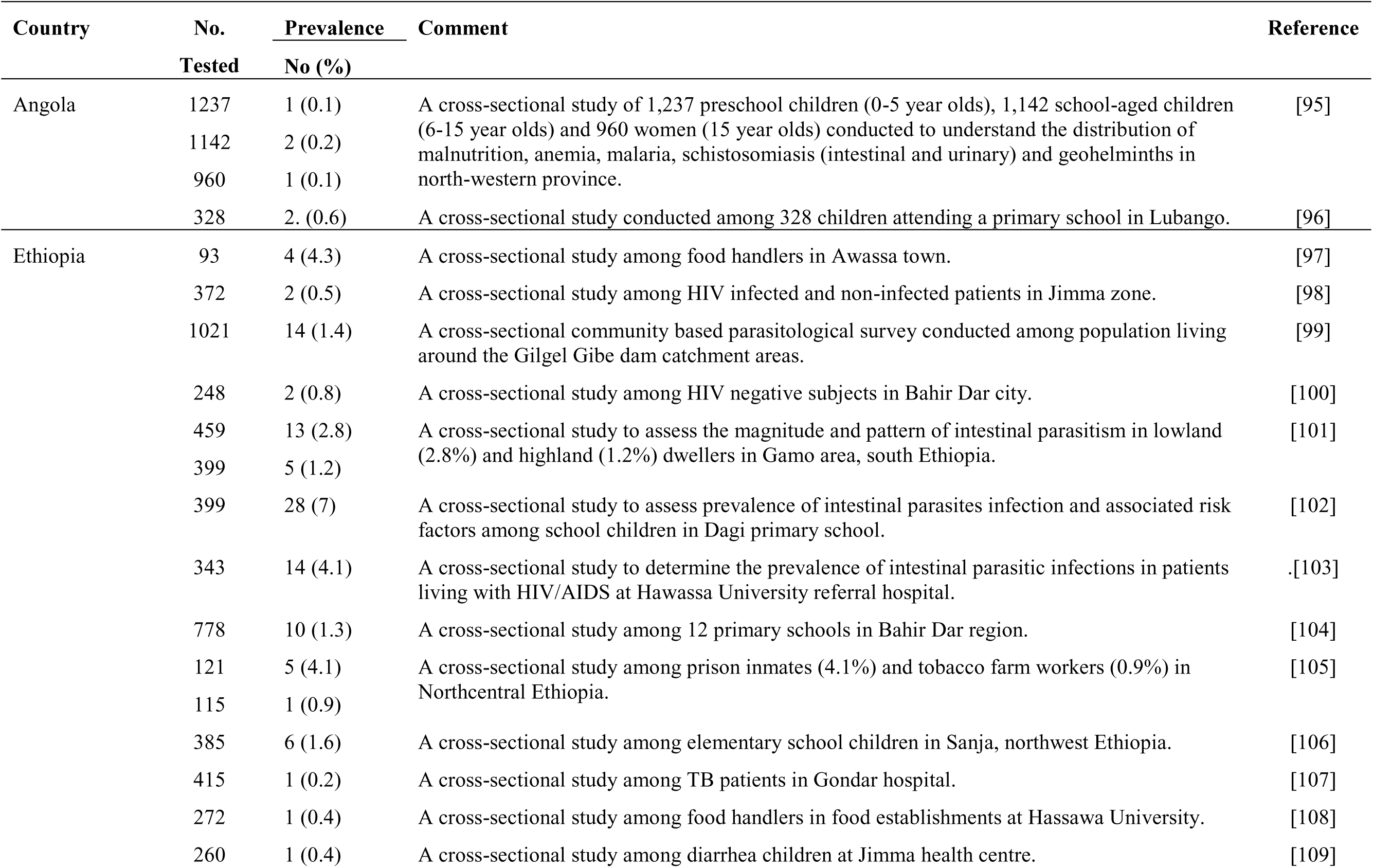
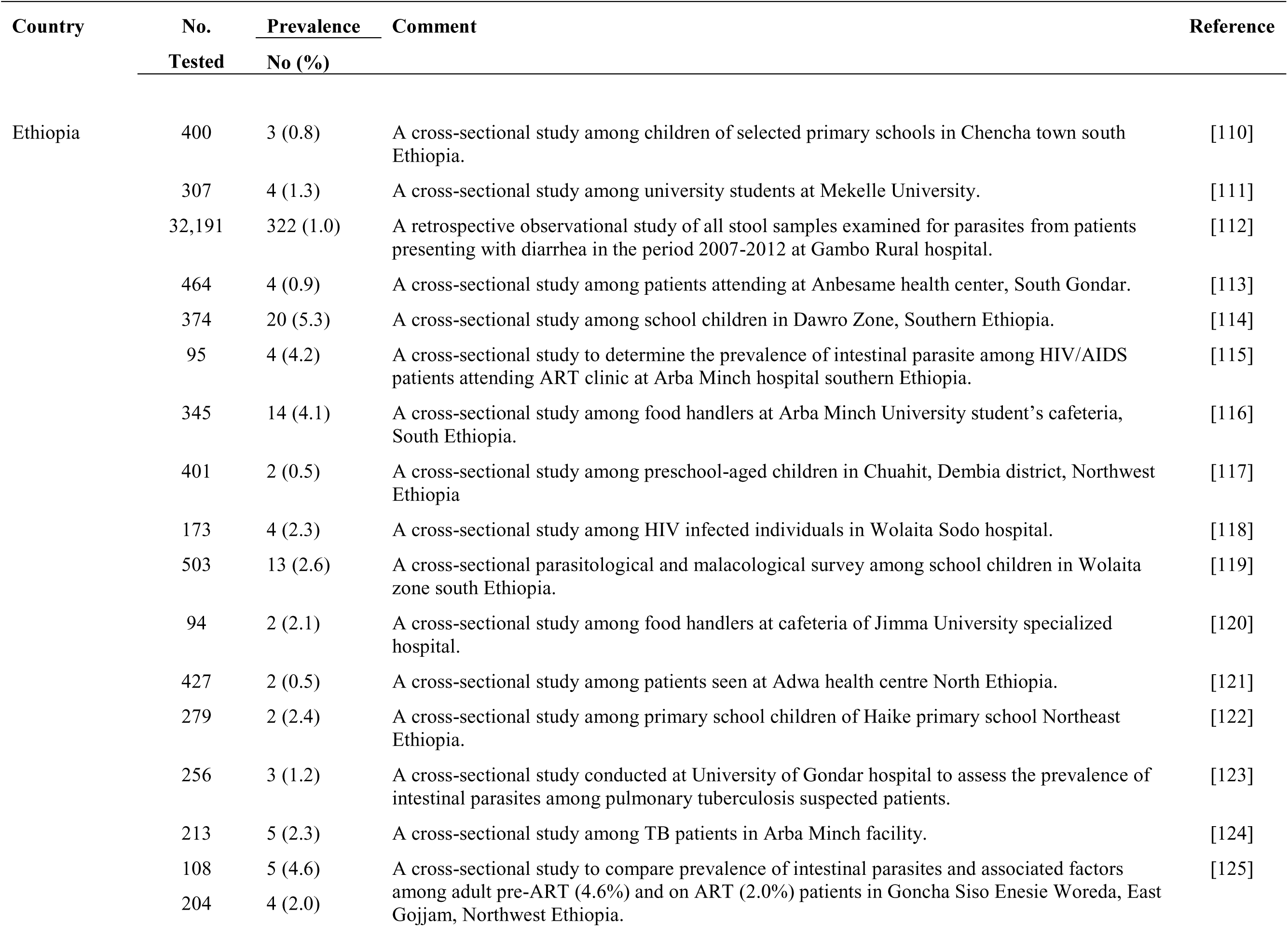
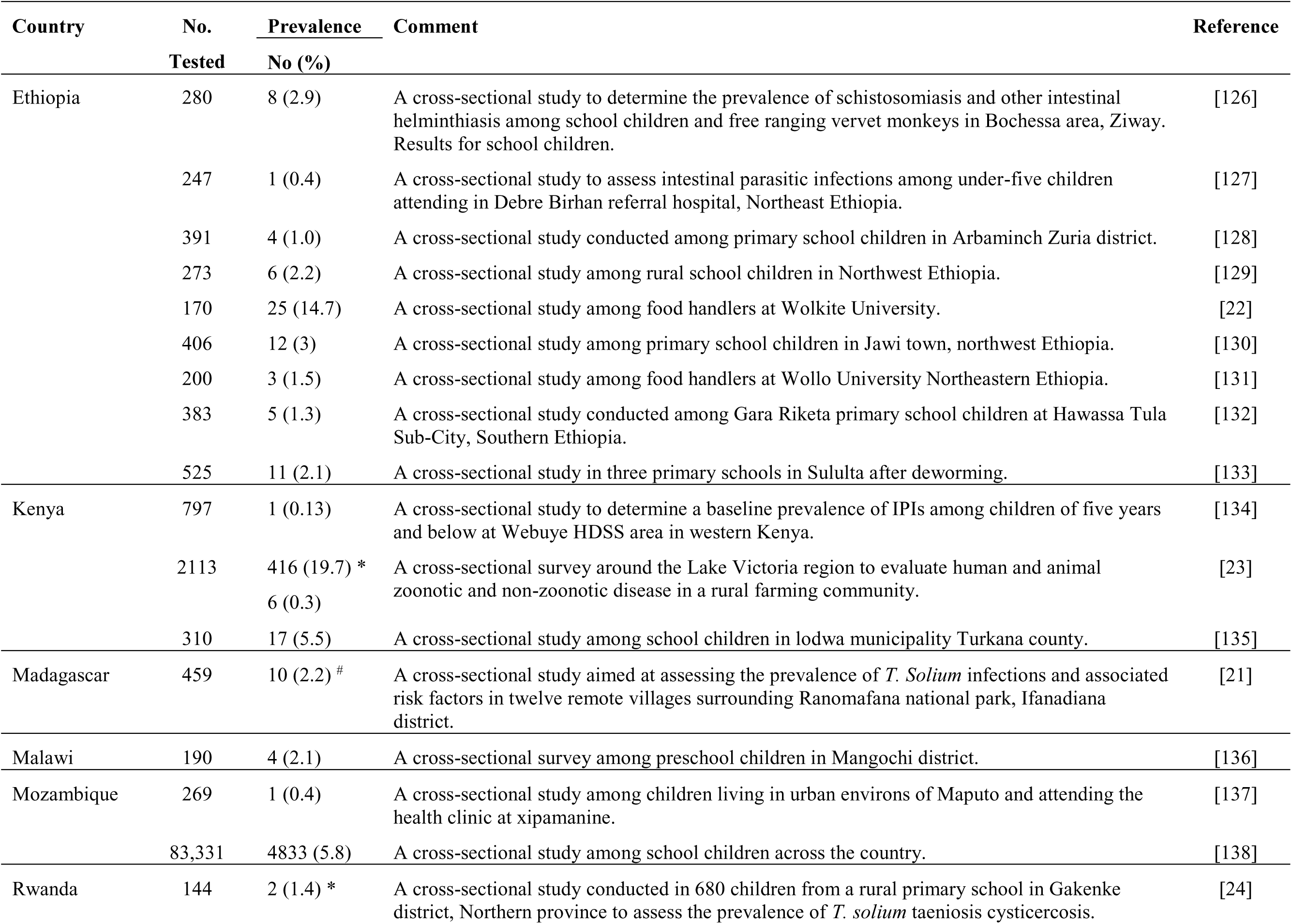
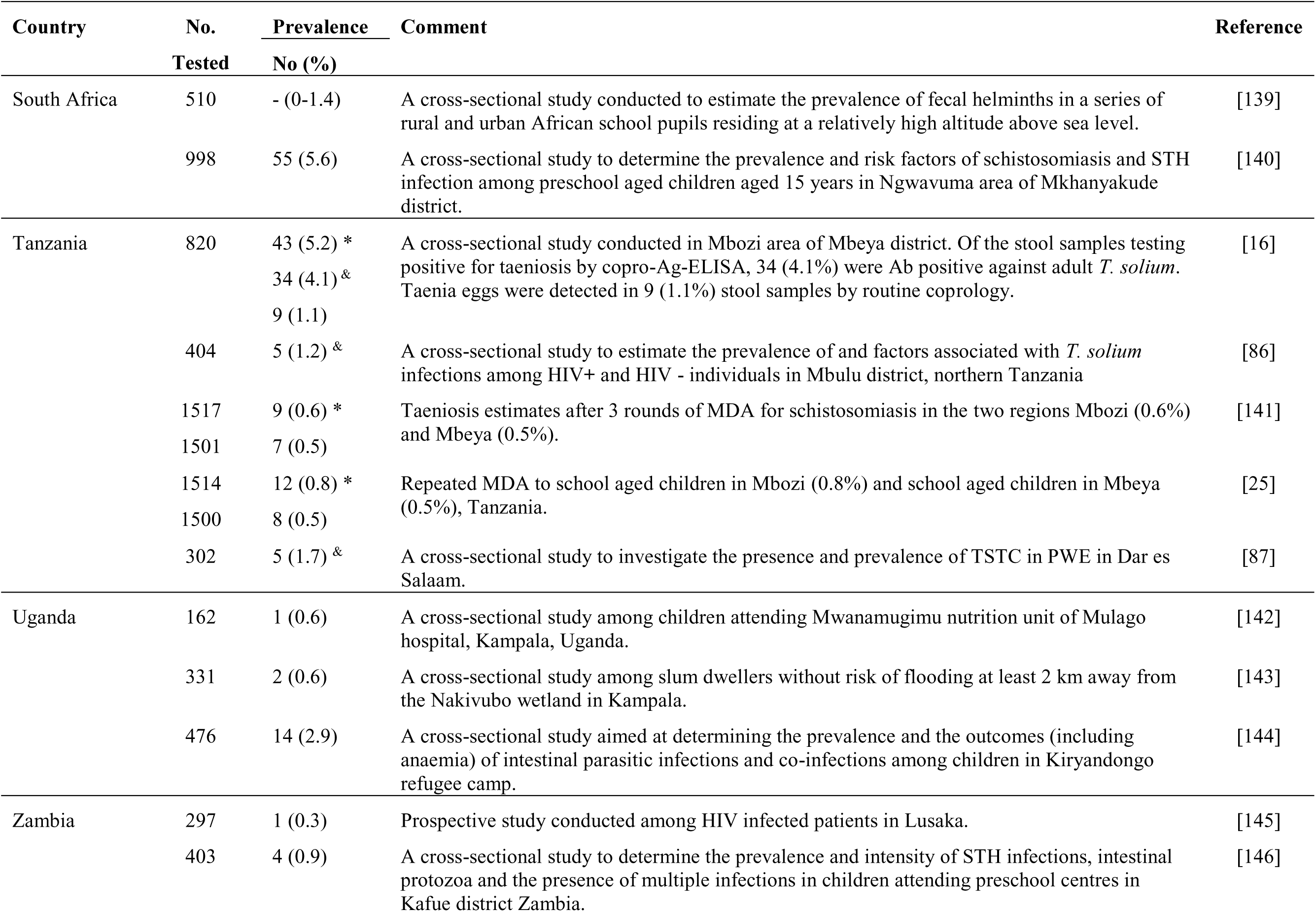
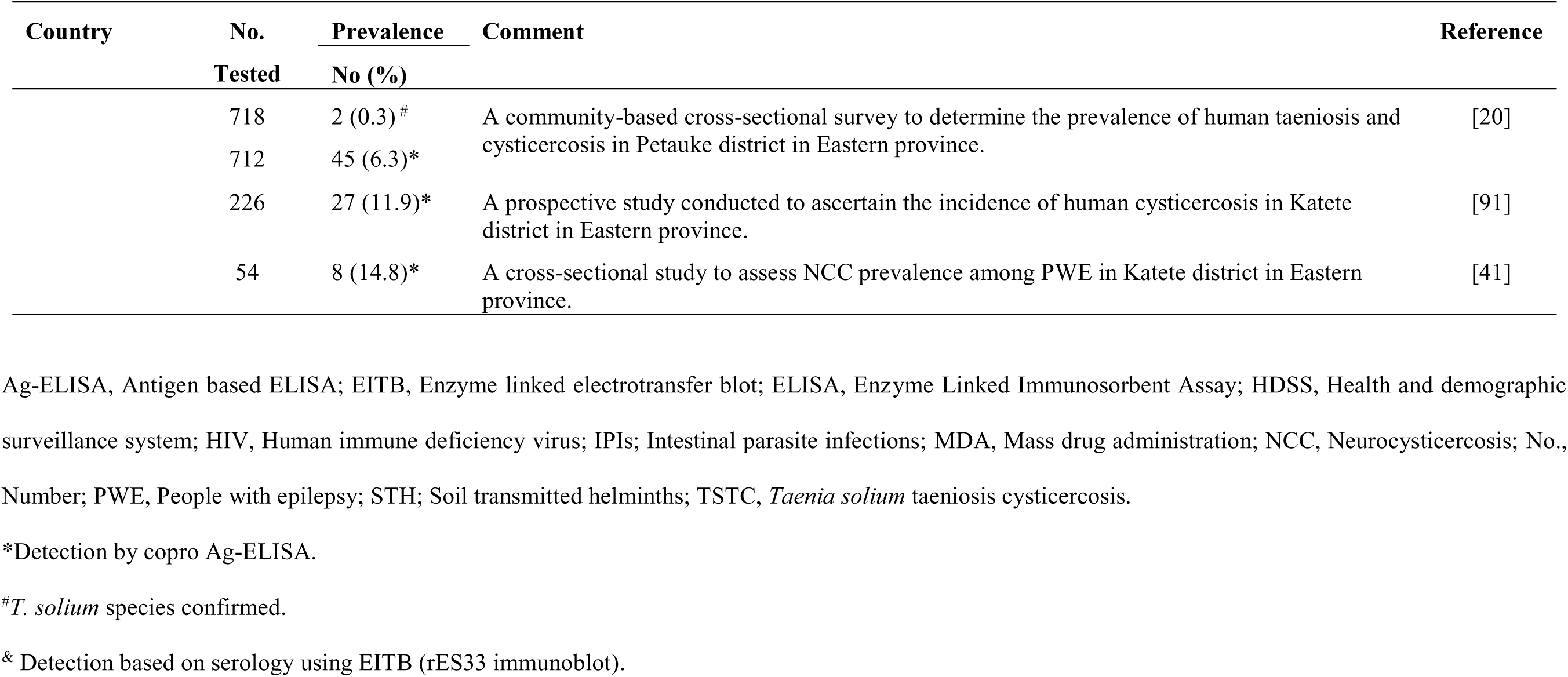
Aggregated human taeniosis cases detected by stool microscopy and reported as Taenia spp in eastern and southern Africa (2000 - 2022).

| Country | No.<br>Tested | Prevalence<br>No (%) | Comment | Reference |
| --- | --- | --- | --- | --- |
| Angola | 1237 | 1 (0.1) | A cross-sectional study of 1,237 preschool children (0-5 year olds), 1,142 school-aged children (6-15 year olds) and 960 women (15 year olds) conducted to understand the distribution of malnutrition, anemia, malaria, schistosomiasis (intestinal and urinary) and geohelminths in north-western province. | [95] |
|  | 1142 | 2 (0.2) |  |  |
|  | 960 | 1 (0.1) |  |  |
|  | 328 | 2. (0.6) | A cross-sectional study conducted among 328 children attending a primary school in Lubango. | [96] |
| Ethiopia | 93 | 4 (4.3) | A cross-sectional study among food handlers in Awassa town. | [97] |
|  | 372 | 2 (0.5) | A cross-sectional study among HIV infected and non-infected patients in Jimma zone. | [98] |
|  | 1021 | 14 (1.4) | A cross-sectional community based parasitological survey conducted among population living around the Gilgel Gibe dam catchment areas. | [99] |
|  | 248 | 2 (0.8) | A cross-sectional study among HIV negative subjects in Bahir Dar city. | [100] |
|  | 459 | 13 (2.8) | A cross-sectional study to assess the magnitude and pattern of intestinal parasitism in lowland (2.8%) and highland (1.2%) dwellers in Gamo area, south Ethiopia. | [101] |
|  | 399 | 5 (1.2) |  |  |
|  | 399 | 28 (7) | A cross-sectional study to assess prevalence of intestinal parasites infection and associated risk factors among school children in Dagi primary school. | [102] |
|  | 343 | 14 (4.1) | A cross-sectional study to determine the prevalence of intestinal parasitic infections in patients living with HIV/AIDS at Hawassa University referral hospital. | [103] |
|  | 778 | 10 (1.3) | A cross-sectional study among 12 primary schools in Bahir Dar region. | [104] |
|  | 121 | 5 (4.1) | A cross-sectional study among prison inmates (4.1%) and tobacco farm workers (0.9%) in Northcentral Ethiopia. | [105] |
|  | 115 | 1 (0.9) |  |  |
|  | 385 | 6 (1.6) | A cross-sectional study among elementary school children in Sanja, northwest Ethiopia. | [106] |
|  | 415 | 1 (0.2) | A cross-sectional study among TB patients in Gondar hospital. | [107] |
|  | 272 | 1 (0.4) | A cross-sectional study among food handlers in food establishments at Hassawa University. | [108] |
|  | 260 | 1 (0.4) | A cross-sectional study among diarrhea children at Jimma health centre. | [109] |

| Country | No. | Prevalence | Comment | Reference |
| --- | --- | --- | --- | --- |
|  | Tested | No (%) |  |  |
| Ethiopia | 400 | 3 (0.8) | A cross-sectional study among children of selected primary schools in Chenchu town south Ethiopia. | [110] |
|  | 307 | 4 (1.3) | A cross-sectional study among university students at Mekelle University. | [111] |
|  | 32,191 | 322 (1.0) | A retrospective observational study of all stool samples examined for parasites from patients presenting with diarrhea in the period 2007-2012 at Gambo Rural hospital. | [112] |
|  | 464 | 4 (0.9) | A cross-sectional study among patients attending at Anbesame health center, South Gondar. | [113] |
|  | 374 | 20 (5.3) | A cross-sectional study among school children in Dawro Zone, Southern Ethiopia. | [114] |
|  | 95 | 4 (4.2) | A cross-sectional study to determine the prevalence of intestinal parasite among HIV/AIDS patients attending ART clinic at Arba Minch hospital southern Ethiopia. | [115] |
|  | 345 | 14 (4.1) | A cross-sectional study among food handlers at Arba Minch University student's cafeteria, South Ethiopia. | [116] |
|  | 401 | 2 (0.5) | A cross-sectional study among preschool-aged children in Chuahit, Dembia district, Northwest Ethiopia | [117] |
|  | 173 | 4 (2.3) | A cross-sectional study among HIV infected individuals in Wolaita Sodo hospital. | [118] |
|  | 503 | 13 (2.6) | A cross-sectional parasitological and malacological survey among school children in Wolaita zone south Ethiopia. | [119] |
|  | 94 | 2 (2.1) | A cross-sectional study among food handlers at cafeteria of Jimma University specialized hospital. | [120] |
|  | 427 | 2 (0.5) | A cross-sectional study among patients seen at Adwa health centre North Ethiopia. | [121] |
|  | 279 | 2 (2.4) | A cross-sectional study among primary school children of Haike primary school Northeast Ethiopia. | [122] |
|  | 256 | 3 (1.2) | A cross-sectional study conducted at University of Gondar hospital to assess the prevalence of intestinal parasites among pulmonary tuberculosis suspected patients. | [123] |
|  | 213 | 5 (2.3) | A cross-sectional study among TB patients in Arba Minch facility. | [124] |
|  | 108 | 5 (4.6) | A cross-sectional study to compare prevalence of intestinal parasites and associated factors among adult pre-ART (4.6%) and on ART (2.0%) patients in Goncha Siso Enesie Woreda, East Gojjam, Northwest Ethiopia. | [125] |
|  | 204 | 4 (2.0) |  |  |
| Ethiopia | 280 | 8 (2.9) | A cross-sectional study to determine the prevalence of schistosomiasis and other intestinal helminthiasis among school children and free ranging vervet monkeys in Bochessa area, Ziway. Results for school children. | [126] |
|  | 247 | 1 (0.4) | A cross-sectional study to assess intestinal parasitic infections among under-five children attending in Debre Birhan referral hospital, Northeast Ethiopia. | [127] |
|  | 391 | 4 (1.0) | A cross-sectional study conducted among primary school children in Arbaminch Zuria district. | [128] |
|  | 273 | 6 (2.2) | A cross-sectional study among rural school children in Northwest Ethiopia. | [129] |
|  | 170 | 25 (14.7) | A cross-sectional study among food handlers at Wolkite University. | [22] |
|  | 406 | 12 (3) | A cross-sectional study among primary school children in Jawi town, northwest Ethiopia. | [130] |
|  | 200 | 3 (1.5) | A cross-sectional study among food handlers at Wollo University Northeastern Ethiopia. | [131] |
|  | 383 | 5 (1.3) | A cross-sectional study conducted among Gara Riketa primary school children at Hawassa Tula Sub-City, Southern Ethiopia. | [132] |
|  | 525 | 11 (2.1) | A cross-sectional study in three primary schools in Sululta after deworming. | [133] |
| Kenya | 797 | 1 (0.13) | A cross-sectional study to determine a baseline prevalence of IPIs among children of five years and below at Webuye HDSS area in western Kenya. | [134] |
|  | 2113 | 416 (19.7) *<br>6 (0.3) | A cross-sectional survey around the Lake Victoria region to evaluate human and animal zoonotic and non-zoonotic disease in a rural farming community. | [23] |
|  | 310 | 17 (5.5) | A cross-sectional study among school children in lodwa municipality Turkana county. | [135] |
| Madagascar | 459 | 10 (2.2) # | A cross-sectional study aimed at assessing the prevalence of *T. Solium* infections and associated risk factors in twelve remote villages surrounding Ranomafana national park, Ifanadiana district. | [21] |
| Malawi | 190 | 4 (2.1) | A cross-sectional survey among preschool children in Mangochi district. | [136] |
| Mozambique | 269 | 1 (0.4) | A cross-sectional study among children living in urban environs of Maputo and attending the health clinic at xipamanine. | [137] |
|  | 83,331 | 4833 (5.8) | A cross-sectional study among school children across the country. | [138] |
| Rwanda | 144 | 2 (1.4) * | A cross-sectional study conducted in 680 children from a rural primary school in Gakenke district, Northern province to assess the prevalence of *T. solium* taeniosis cysticercosis. | [24] |
| South Africa | 510 | - (0-1.4) | A cross-sectional study conducted to estimate the prevalence of fecal helminths in a series of rural and urban African school pupils residing at a relatively high altitude above sea level. | [139] |
|  | 998 | 55 (5.6) | A cross-sectional study to determine the prevalence and risk factors of schistosomiasis and STH infection among preschool aged children aged 15 years in Ngwavuma area of Mkhanyakude district. | [140] |
| Tanzania | 820 | 43 (5.2) *<br>34 (4.1) &<br>9 (1.1) | A cross-sectional study conducted in Mbozi area of Mbeya district. Of the stool samples testing positive for taeniosis by copro-Ag-ELISA, 34 (4.1%) were Ab positive against adult *T. solium*. Taenia eggs were detected in 9 (1.1%) stool samples by routine coprology. | [16] |
|  | 404 | 5 (1.2) & | A cross-sectional study to estimate the prevalence of and factors associated with *T. solium* infections among HIV+ and HIV - individuals in Mbulu district, northern Tanzania | [86] |
|  | 1517 | 9 (0.6) * | Taeniosis estimates after 3 rounds of MDA for schistosomiasis in the two regions Mbozi (0.6%) and Mbeya (0.5%). | [141] |
|  | 1501 | 7 (0.5) |  |  |
|  | 1514 | 12 (0.8) * | Repeated MDA to school aged children in Mbozi (0.8%) and school aged children in Mbeya (0.5%), Tanzania. | [25] |
|  | 1500 | 8 (0.5) |  |  |
|  | 302 | 5 (1.7) & | A cross-sectional study to investigate the presence and prevalence of TSTC in PWE in Dar es Salaam. | [87] |
| Uganda | 162 | 1 (0.6) | A cross-sectional study among children attending Mwanamugimu nutrition unit of Mulago hospital, Kampala, Uganda. | [142] |
|  | 331 | 2 (0.6) | A cross-sectional study among slum dwellers without risk of flooding at least 2 km away from the Nakivubo wetland in Kampala. | [143] |
|  | 476 | 14 (2.9) | A cross-sectional study aimed at determining the prevalence and the outcomes (including anaemia) of intestinal parasitic infections and co-infections among children in Kiryandongo refugee camp. | [144] |
| Zambia | 297 | 1 (0.3) | Prospective study conducted among HIV infected patients in Lusaka. | [145] |
|  | 403 | 4 (0.9) | A cross-sectional study to determine the prevalence and intensity of STH infections, intestinal protozoa and the presence of multiple infections in children attending preschool centres in Kafue district Zambia. | [146] |
|  | 718 | 2 (0.3) <sup>#</sup> | A community-based cross-sectional survey to determine the prevalence of human taeniosis and cysticercosis in Petauke district in Eastern province. | [20] |
|  | 712 | 45 (6.3)* |  |  |
|  | 226 | 27 (11.9)* | A prospective study conducted to ascertain the incidence of human cysticercosis in Katete district in Eastern province. | [91] |
|  | 54 | 8 (14.8)* | A cross-sectional study to assess NCC prevalence among PWE in Katete district in Eastern province. | [41] |

Ag-ELISA, Antigen based ELISA; EITB, Enzyme linked electrotransfer blot; ELISA, Enzyme Linked Immunosorbent Assay; HDSS, Health and demographic surveillance system; HIV, Human immune deficiency virus; IPIs; Intestinal parasite infections; MDA, Mass drug administration; NCC, Neurocysticercosis; No., Number; PWE, People with epilepsy; STH; Soil transmitted helminths; TSTC, *Taenia solium* taeniosis cysticercosis.
\*Detection by copro Ag-ELISA.
<sup>#</sup>*T. solium* species confirmed.
& Detection based on serology using EITB (rES33 immunoblot).

Several groups of people were studied for human taeniosis with 25 records reporting taeniosis cases in school children, 18 records on patients presenting at health facilities, 15 on consenting community volunteers, and six on food handlers. There was great variation in aggregated numbers of human taeniosis cases across countries with 595 cases reported from Ethiopia alone and even more (4834) reported from Mozambique. The number of human taeniosis cases identified for the other nine countries in ESA is shown in (Fig 8).

**Fig 8.**
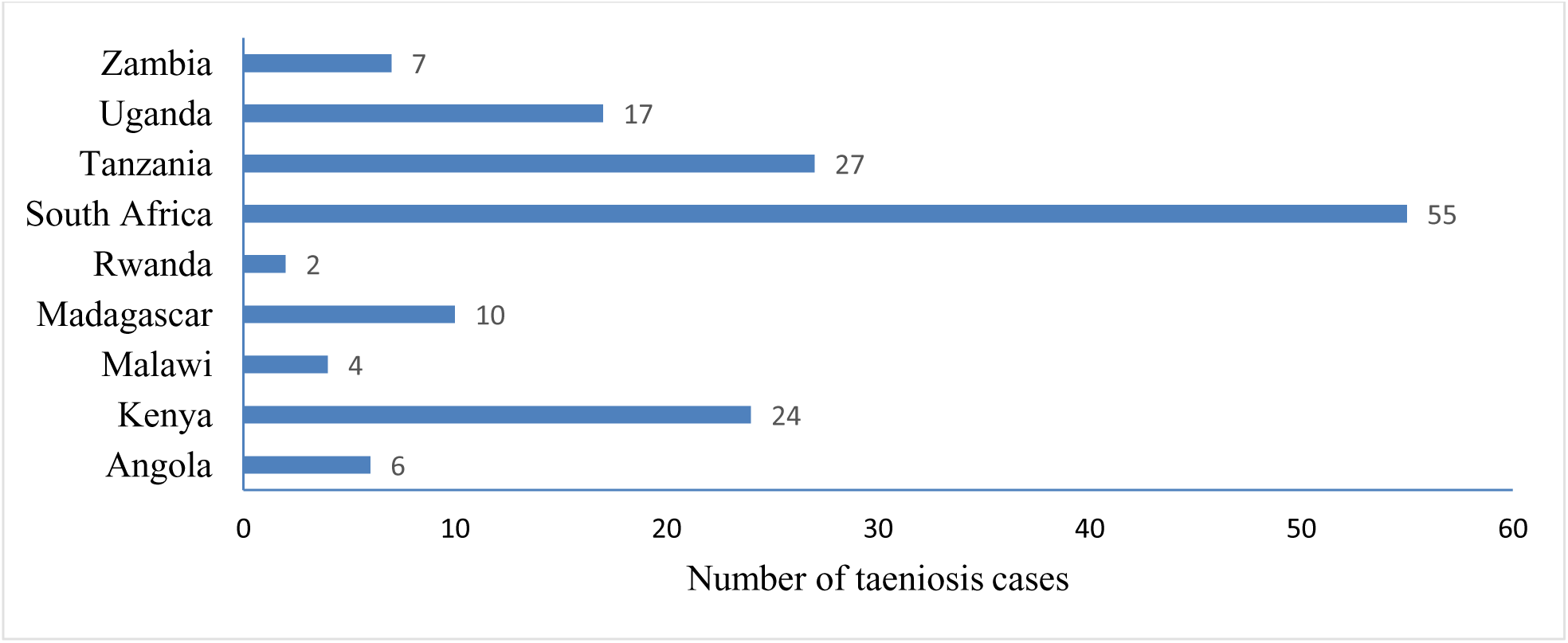
Taeniosis cases reported in eastern and southern Africa between 2000 – 2020

The largest number of cases were reported as *Taenia spp.* (Table 6) without specifying the type of species. In only two records from Madagascar and Zambia, *T. solium* was confirmed using PCR [20, 21]. Microscopy only was employed for 53 studies, and copro Ag-ELISA only in three studies. Five studies used both copro Ag-ELISA and microscopy for taeniosis diagnosis. Three studies used the rES33 EITB for the presence of a tapeworm antibodies in the body. Immunological data (copro Ag-ELISA or rES33 EITB) on taeniosis was only available from Kenya, Rwanda, Tanzania, and Zambia.

Within the ESA region, the prevalence of human taeniosis based on microscopy ranged between 0.1 - 14.7% with the highest prevalence reported in a study conducted among food handlers at Wolkite University in Ethiopia [22] (Figs 7 and 9). Among studies based on copro Ag-ELISA conducted in Kenya, Rwanda, Tanzania, and Zambia, the highest prevalence was reported in Kenya at 19.7% [23]. Rwanda had a taeniosis prevalence of 1.4% [24], Tanzania ranged between 0.5 to 5.2% [16, 25] [16], [22] and Zambia ranged between 0.3 to 13.8% [20, 26] (Fig 10). For details on sources refer to Table 6.

**Fig 9.**
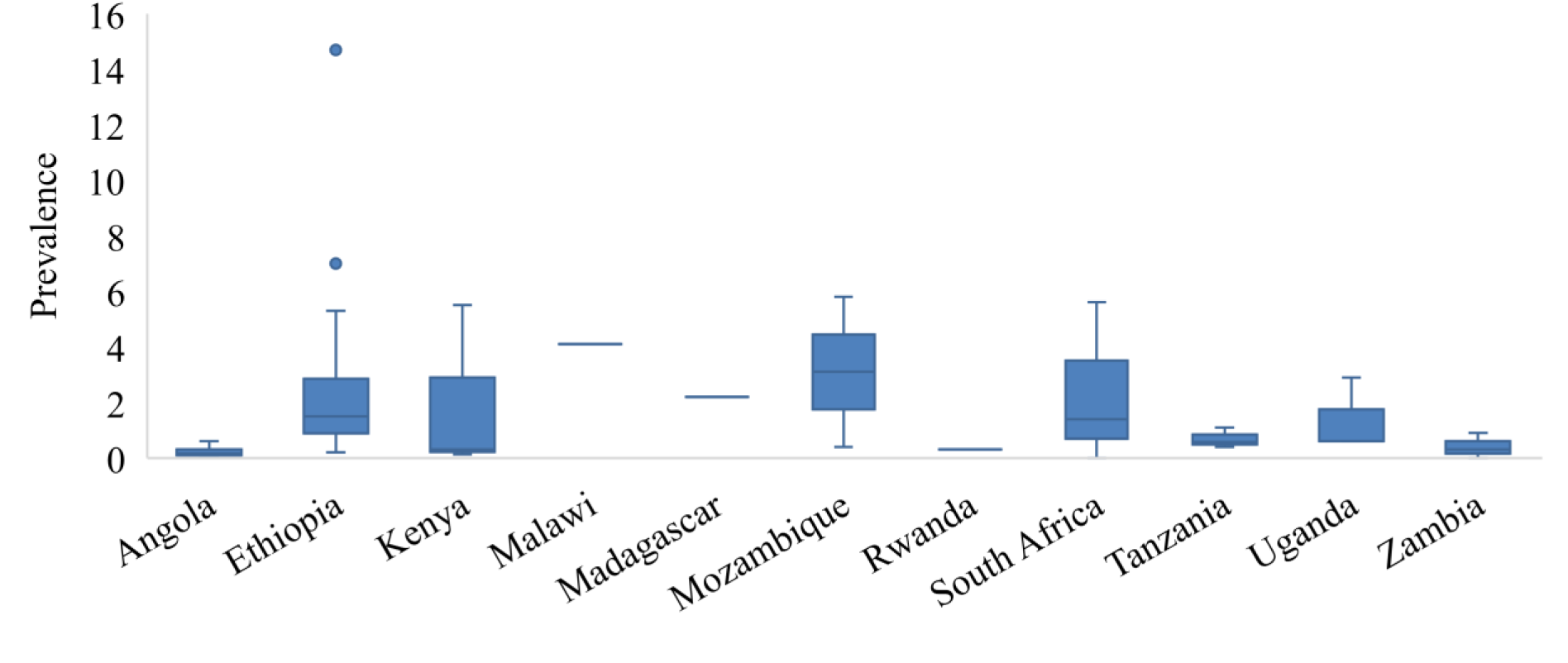
Prevalence of taeniosis in eastern and southern Africa based on microscopy data in reviewed studies

**Fig 10.**
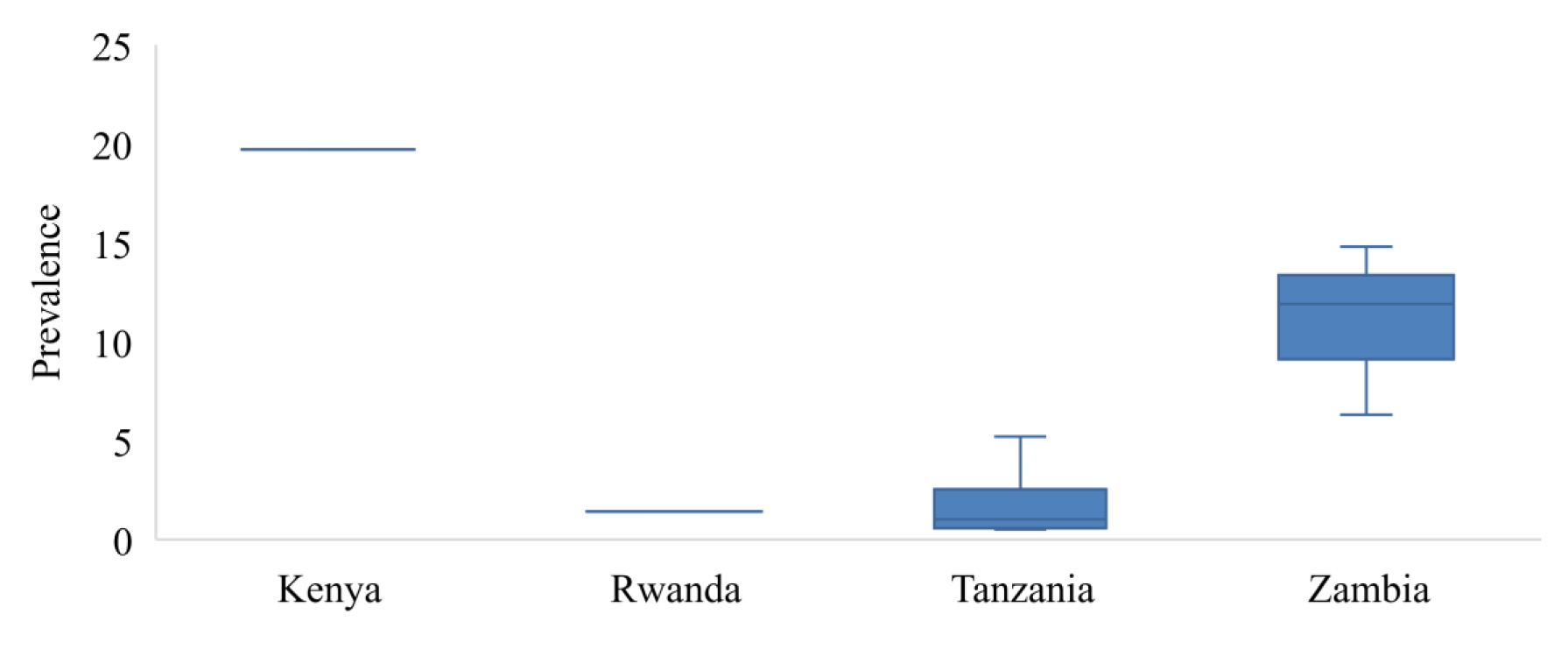
Prevalence of taeniosis based on copro-Ag ELISA in reviewed studies in eastern and southern Africa.

## Discussion

This study aimed at collecting epidemiological data on *T. solium* infections in humans in the ESA region for the period 2000 to 2022. For both cysticercosis and taeniosis, there was no data from Comoros, Djibouti, Eritrea, Mayotte, Reunion, Seychelles, Socotra, Somalia and Somaliland that could be retrieved. Somalia has for some time now experienced internal conflicts and this may have a bearing on the diagnosis and reporting of human cysticercosis and taeniosis cases. For the other six island states and territories, a lack of scientific interest in reporting cases has been cited [27]. The same can be said about Botswana and Lesotho from which data on cysticercosis and taeniosis was also lacking. Cysticercosis data was lacking in even more countries.

According to the World Bank IBRD-IDA report of 2022 [28] the population using safely managed sanitation services for both Comoros and Djibouti only stood at 37% with no data for other island states. People practicing open defecation were higher in Eritrea (67%) with Somalia reporting (23%) and Djibouti (16%). On the Réunion island, pork is said to be the most popular meat with a total annual pork consumption of about 20 000 tones. Half of this is produced on the island and the rest is imported frozen from Europe [29]. Comoros on the other hand has a small livestock sector with most of its meat including pork imported from Tanzania, Madagascar and France [30]. Seventy per cent of the Somali human population subsists in pastoralism with sheep and goats being the dominant animals. Camels are primarily raised for milk production and small ruminants are for generating cash income for the family [31]. For Mayotte, Eritrea and Djibouti, information on pig rearing could not be found as these countries focus mostly on cattle, sheep, goats, and camels [32]. However, for Eritrea, pig breeding has recently begun to be promoted [33] and so is the case for Seychelles whose 2011 census of agriculture showed that 2% (483/24770) of the households were raising pigs in small holdings [34]. Therefore, while it is not possible to ascertain the presence or absence of human *T. solium* infections in these countries, there is a possibility that these infections do exist as pork consumption seems to be practiced and risk factors are present; some countries even import pork from confirmed *T. solium* endemic countries. Therefore, the numbers collected for this review represent an underestimation of the *T. solium* epidemiology for human disease in ESA.

Regarding human cysticercosis cases, tapeworm carriers are the sole cause of infection in both rural and urban areas of the ESA region [10, 35]. Tapeworm carriers, if not treated, pose a risk to themselves if they ingest infective eggs leading to cysticercosis but can also pose a risk to other people in contact [21, 36]. A study in Madagascar found at a household level, the overall seroprevalence of IgG detected by Ab-ELISA and EITB within the same household in which a tapeworm carrier has been detected was 46% and 34% respectively [21]. Individual human cysticercosis cases were identified from a third of the 27 ESA countries. South Africa reported the highest number of individual human cysticercosis cases followed by Madagascar. This could be due to increased awareness of cysticercosis as a possible diagnosis in, for example, people with epilepsy in these countries compared to other countries within the region. South Africa also has more resources compared to the other countries within the region. For example, as early as 2003 South Africa already had 214 CT scans and 111 neurologists. This is an advantage in terms of diagnosis and management of particularly NCC that is not present in other countries within the region [37]. With poor pig management, poor meat inspection, and poor sanitation those who do not eat pork are equally at risk of infection with cysticercosis as those eating pork [38–40]. Additionally, certain cultural practices within the region have also been cited as contributing factors to cysticercosis. For example, in South Africa, it has been reported that some unqualified traditional healers “baloi” used *Taenia* segments and added them to medicinal mixtures as strengthening ingredients. There are also reports of *T. solium* segments being added to the beer of unfaithful husbands or lovers as punishment [37].

NCC has been associated with up to 57% of epilepsy cases in sub–Saharan Africa to which the region under consideration belongs [7, 41]. This is higher than the global average of 30% of NCC among PWE [6]. This association is higher than the global average of 30% [6]. For most countries within ESA, the diagnosis of NCC is problematic due to the scarcity of neuroimaging, serology and trained neurologists [42, 43]. This diagnostic challenge is not only present in ESA but also in Europe where a lack of awareness of NCC leads to underdiagnoses [44]. Only a quarter of the countries within the ESA reported data on NCC and epilepsy. Epilepsy is one of the most common neurological disorders in many parts of sub-Saharan Africa and NCC seems to be a major cause of it in *T. solium* cysticercosis endemic areas. Diagnosing NCC among people with epilepsy is vital to prevent further morbidity and mortality from the disease as well as to reduce the negative socio-cultural beliefs associated with epilepsy in the region [42,45–47].

Regarding human *T. solium* taeniosis in ESA, only Kenya, Madagascar, Rwanda, Tanzania, and Zambia conducted community-based studies and employed serological tests that specifically aimed at determining the burden of infection. The other countries reported data on taeniosis as incidental findings following microscopic examinations of stool samples conducted for soil-transmitted helminths. The reported human taeniosis cases were also reported on an aggregated level without evidence of species determination. Thus, cases reported as either taeniosis or *taenia spp* could not for instance be differentiated from cases due to *T. saginata* which is also widely distributed in ESA [27]. Only two studies, one from Madagascar and another from Zambia were able to confirm the *T. solium* species after collecting tapeworm proglottids [20, 21].

This lack of species differentiation creates uncertainty regarding the epidemiology of *T. solium* infections in ESA. There is therefore a possible underestimation of the magnitude of *T. solium* taeniosis in the region. This is worsened by the fact that *T. solium* taeniosis is not even a notifiable disease in most of these countries. Application of molecular methods to differentiate *T. solium* species on stool examination is not widely practiced and it is difficult to differentiate *taenia* eggs based on microscopy [35, 48]. More sensitive and specific diagnostic tools need to be used to identify the true prevalence and outline the epidemiology of *T. solium*.

Human taeniosis in the region is due to the high prevalence of porcine cysticercosis in ESA countries which is ranked amongst the highest in the world [10]. Over the last decade, pig production as a risk factor for human taeniosis in the rural communities of ESA countries has increased mainly because pig raising is an attractive alternative to other animals as it does not require grazing land compared to ruminants. Free-range pig keeping is popular, especially in rural areas of the region as it is easy and cheap. This coupled with the general lack of slaughterhouses and inspection of pork with a lack of sanitation facilities has contributed to the risk for people to attract taeniosis and cysticercosis [3,10,49].

### Study limitation

Our study is limited by the fact that search platforms were used only for online publications. Thus, unpublished data which could have contributed significantly to a more nuanced picture of the epidemiology of human *T. solium* infections in ESA could have been missed. Another limitation of our study is the difficulty in comparing epidemiological data from different study populations using different diagnostic tests used and the different diagnostic approaches.

## Conclusion

In this review on human *T. solium* taeniosis and (neuro) cysticercosis within ESA, we found wide variations in the prevailing prevalent estimates depending on the quality of the study and the diagnostic methods used. There remain large gaps with regards to *T. solium* infections within the ESA region with 11 countries without any information on human taeniosis and cysticercosis, and even more countries (18) without any information for cysticercosis.

Considering the public health and economic impact that *T. solium* infections have, it is important to understand its epidemiology in humans. Human taeniosis, cysticercosis and NCC cases need to be reported and notified to public health authorities. More community-based surveys as well as ante-mortem and postmortem porcine surveys must be conducted for control and surveillance purposes. Lack of awareness about the presence of *T. solium* among medical, community and government authorities including inadequate technology for diagnosis of taeniosis, cysticercosis and NCC, in both humans and pigs (cysticercosis) remains a challenge that needs to be addressed for proper surveillance, prevention and control.

## Data Availability

All references found eligible in our review are included in the supplementary materials (S2 Table).

## Acknowledgment

None.

## Supplement

**S1 File. Documentation of literature search. (DOCX)**

**S2 File. Data collection forms. (DOCX)**

**S1 Table. PRISMA 2020 checklist. (DOCX)**

**S2 Table. References retrieved through online international databases. (XLS)**

## Availability of data and material

All references found eligible in our review are included in the supplementary materials (S2 Table).

## Author contributions

GZ, CM and HS conducted the systematic review of the literature and extracted data. GZ analysed the data and drafted the first version of the manuscript. AA, ASW, CM, DS, GZ, IGK, KEM, VS contributed to the design of the study. All authors contributed to interpretation of data and writing of the paper. All authors read and approved the final manuscript.

## References

[1] J. Sotelo, O.H. Del Brutto, Review of neurocysticercosis, Neurosurg. Focus. 12 (2002) e1. https://doi.org/10.3171/foc.2002.12.6.2.

[2] K.D. Murrell, WHO/FAO/OIE guidelines for the surveillance, prevention and control of taeniosis/cysticercosis. Paris: World Health Organisation for Animal Health (OIE), 2005, 2005. http://www.oie.int (accessed May 6, 2019).

[3] H. Carabin, A. Millogo, A. Cissé, S. Gabriël, I. Sahlu, P. Dorny, C. Bauer, Z. Tarnagda, L.D. Cowan, R. Ganaba, Prevalence of and Factors Associated with Human Cysticercosis in 60 Villages in Three Provinces of Burkina Faso, PLoS Negl. Trop. Dis. 9 (2015) e0004248. https://doi.org/10.1371/journal.pntd.0004248.

[4] A. Winkler, H. Richter, Landscape analysis : management of neurocysticercosis with an emphasis on low- and middle-income countries, World Heal. Organ. (2015) 65. www.neurokopfzentrum.med.tum.de/neurologie/42b.html (accessed February 22, 2018).

[5] D. Stelzle, V. Schmidt, L. Keller, B.J. Ngowi, W. Matuja, G. Escheu, P. Hauke, V. Richter, E. Ovuga, B. Pfausler, E. Schmutzhard, A. Amos, W. Harrison, J. Kaducu, A.S. Winkler, Characteristics of people with epilepsy and Neurocysticercosis in three eastern African countries–A pooled analysis, PLoS Negl. Trop. Dis. 16 (2022) e0010870. https://doi.org/10.1371/journal.pntd.0010870.

[6] P.C. Ndimubanzi, H. Carabin, C.M. Budke, H. Nguyen, Y.J. Qian, E. Rainwater, M. Dickey, S. Reynolds, J.A. Stoner, A systematic review of the frequency of neurocyticercosis with a focus on people with epilepsy, PLoS Negl. Trop. Dis. 4 (2010) e870. https://doi.org/10.1371/journal.pntd.0000870.

[7] L.F. Owolabi, B. Adamu, A.M. Jibo, S.D. Owolabi, A.I. Imam, I.D. Alhaji, Neurocysticercosis in people with epilepsy in Sub-Saharan Africa: A systematic review and meta-analysis of the prevalence and strength of association, SeizureEuropean J. Epilepsy. 76 (2020) 1–11. https://doi.org/10.1016/j.seizure.2020.01.005.

[8] E. Bruno, A. Bartoloni, L. Zammarchi, M. Strohmeyer, F. Bartalesi, J. Bustos, S. Santivañez, H.H. Garcia, A. Nicoletti, M. Bonati, F. Severino, V. Confalonieri, C. Pandolfini, Z. Bisoffi, D. Buonfrate, A. Angheben, M. Albonico, J. Muñoz, R. Pool, A. Requena-Mendez, M. Roura, A. Hardon, C. Pell, P. Chiodini, J. Moreira, R. Sempértegui, M. Anselmi, E. Gotuzzo, M.A. Mena, F. Torrico, D. Lozano, G.C. Rojas, T.H. Cabrera, J.O. Morón, I.A. Cuellar, J.A. Suarez, G. Tognoni, C.L. Caro, Epilepsy and Neurocysticercosis in Latin America: A Systematic Review and Meta-analysis, PLoS Negl. Trop. Dis. 7 (2013). https://doi.org/10.1371/journal.pntd.0002480.

[9] D. Ng-Nguyen, R.J. Traub, V.A.T. Nguyen, K. Breen, M.A. Stevenson, Spatial distribution of Taenia solium exposure in humans and pigs in the Central Highlands of Vietnam, PLoS Negl. Trop. Dis. 12 (2018). https://doi.org/10.1371/journal.pntd.0006810.

[10] I.K. Phiri, H. Ngowi, S. Afonso, E. Matenga, M. Boa, S. Mukaratirwa, S. Githigia, M. Saimo, C. Sikasunge, N. Maingi, G.W. Lubega, A. Kassuku, L. Michael, S. Siziya, R.C. Krecek, E. Noormahomed, M. Vilhena, P. Dorny, A.L. Willingham, The emergence of Taenia solium cysticercosis in Eastern and Southern Africa as a serious agricultural problem and public health risk, in: Acta Trop., Elsevier, 2003: pp. 13–23. https://doi.org/10.1016/S0001-706X(03)00051-2.

[11] M.J. Page, J.E. McKenzie, P.M. Bossuyt, I. Boutron, T.C. Hoffmann, C.D. Mulrow, L. Shamseer, J.M. Tetzlaff, E.A. Akl, S.E. Brennan, R. Chou, J. Glanville, J.M. Grimshaw, A. Hróbjartsson, M.M. Lalu, T. Li, E.W. Loder, E. Mayo-Wilson, S. McDonald, L.A. McGuinness, L.A. Stewart, J. Thomas, A.C. Tricco, V.A. Welch, P. Whiting, D. Moher, The PRISMA 2020 statement: An updated guideline for reporting systematic reviews, BMJ. 372 (2021). https://doi.org/10.1136/bmj.n71.

[12] M. Viswanathan, A. MT, N. Berkman, S. Chang, L. Hartling, M., et al. McPheeters, Assessing the risk of bias of individual studies in systematic reviews of health care interventions - methods guide – chapter | AHRQ effective health care program, (2016). www.effectivehealthcare.ahrq.gov. (accessed September 7, 2018).

[13] M. Mugabe, D. Ruhangaza, E. Hakizimana, Cysticercosis: Report of 10 cases diagnosed at a district hospital in Rwanda, Mod. Pathol. 32 (2019). http://ovidsp.ovid.com/ovidweb.cgi?T=JS&PAGE=reference&D=emexb&NEWS=N&AN=631814441.

[14] S. Sobnach, S.A. Khosa, S. Pather, S. Longhurst, D. Kahn, P.J. Raubenheimer, First case report of pharyngeal cysticercosis, Trans. R. Soc. Trop. Med. Hyg. 103 (2009) 206–208. https://doi.org/10.1016/j.trstmh.2008.08.017.

[15] C. Himwaze, L.A. Mucheleng’anga, V. Telendiy, A. Hamukale, J. Tembo, N. Kapata, F. Ntoumi, A. Zumla, Cardiac cysticercosis and neurocysticercosis in sudden and unexpected community deaths in Lusaka, Zambia: a descriptive medico-legal post-mortem examination study, Int. J. Infect. Dis. 115 (2022) 195–200. https://doi.org/10.1016/j.ijid.2021.11.042.

[16] G. Mwanjali, C. Kihamia, D.V.C. Kakoko, F. Lekule, H. Ngowi, M.V. Johansen, S.M. Thamsborg, A.L. Willingham, Prevalence and Risk Factors Associated with Human Taenia Solium Infections in Mbozi District, Mbeya Region, Tanzania, PLoS Negl. Trop. Dis. 7 (2013) e2102. https://doi.org/10.1371/journal.pntd.0002102.

[17] A. Buda, O. Dean, H.R. Adams, S. Mwanza-Kabaghe, M.J. Potchen, E.G. Mbewe, P.P. Kabundula, S.M. Moghaddam, M. Mweemba, B. Matoka, M.M. Mathews, G.L. Birbeck, D.R. Bearden, Neurocysticercosis Among Zambian Children and Adolescents With Human Immunodeficiency Virus: A Geographic Information Systems Approach, Pediatr. Neurol. 102 (2020) 36–43. https://doi.org/10.1016/j.pediatrneurol.2019.07.017.

[18] M.M. Diaz, D. Sokhi, J. Noh, A.K. Ngugi, F.J. Minja, P. Reddi, E.M. Fèvre, A.-C.L. Meyer, Prevalence of Epilepsy, Human Cysticercosis, and Porcine Cysticercosis in Western Kenya, Am. J. Trop. Med. Hyg. 106 (2022) 1450–1455. https://doi.org/10.4269/ajtmh.21-0594.

[19] G.S. Ocana, J.C.O. Sablon, I.O. Tamayo, L.A. Arena, L.M.S. Ocana, S. Govender, Neurocysticercosis in patients presenting with epilepsy at St. Elizabeth’s Hospital, Lusikisiki, South African Med. J. 99 (2009) 588–591. https://doi.org/10.7196/SAMJ.3358.

[20] K.E. Mwape, I.K. Phiri, N. Praet, J.B. Muma, G. Zulu, P. van den Bossche, R. de Deken, N. Speybroeck, P. Dorny, S. Gabriël, Taenia solium infections in a rural area of Eastern Zambia-A community based study, PLoS Negl. Trop. Dis. 6 (2012) 1–9. https://doi.org/10.1371/journal.pntd.0001594.

[21] A. Rahantamalala, R.L. Rakotoarison, E. Rakotomalala, M. Rakotondrazaka, J. Kiernan, P.M. Castle, L. Hakami, K. Choi, A.S. Rafalimanantsoa, A. Harimanana, P. Wright, S.G. Lapierre, M. Schoenhals, P.M. Small, L.A. Marcos, I. Vigan-Womas, Prevalence and factors associated with human Taenia solium taeniosis and cysticercosis in twelve remote villages of Ranomafana rainforest, Madagascar, PLoS Negl. Trop. Dis. 16 (2022) e0010265. https://doi.org/10.1371/journal.pntd.0010265.

[22] T.A. Bafa, E.M. Sherif, A.H. Hantalo, G.G. Woldeamanuel, Magnitude of enteropathogens and associated factors among apparently healthy food handlers at Wolkite University Student’s Cafeteria, Southern Ethiopia, BMC Res. Notes. 12 (2019) 1–6. https://doi.org/10.1186/s13104-019-4599-z.

[23] E.M. Fèvre, W.A. de Glanville, L.F. Thomas, E.A.J. Cook, S. Kariuki, C.N. Wamae, An integrated study of human and animal infectious disease in the Lake Victoria crescent small-holder crop-livestock production system, Kenya, BMC Infect. Dis. 17 (2017) 1–14. https://doi.org/10.1186/s12879-017-2559-6.

[24] L. Acosta Soto, L.A. Parker, M.J. Irisarri-Gutiérrez, J.A. Bustos, Y. Castillo, E. Perez, C. Muñoz-Antoli, J.G. Esteban, H.H. García, F.J. Bornay-Llinares, Evidence for Transmission of Taenia solium Taeniasis/Cysticercosis in a Rural Area of Northern Rwanda, Front. Vet. Sci. 8 (2021) 237. https://doi.org/10.3389/fvets.2021.645076.

[25] U.C. Braae, P. Magnussen, B. Ndawi, W. Harrison, F. Lekule, M.V. Johansen, Effect of repeated mass drug administration with praziquantel and track and treat of taeniosis cases on the prevalence of taeniosis in Taenia solium endemic rural communities of Tanzania, Acta Trop. 165 (2017) 246–251. https://doi.org/10.1016/j.actatropica.2015.10.012.

[26] K.E. Mwape, I.K. Phiri, N. Praet, P. Dorny, J.B. Muma, G. Zulu, N. Speybroeck, S. Gabriël, Study and ranking of determinants of taenia solium infections by classification tree models, Am. J. Trop. Med. Hyg. 92 (2015) 56–63. https://doi.org/10.4269/ajtmh.13-0593.

[27] V. Dermauw, P. Dorny, U.C. Braae, B. Devleesschauwer, L.J. Robertson, A. Saratsis, L.F. Thomas, Epidemiology of Taenia saginata taeniosis/cysticercosis: A systematic review of the distribution in southern and eastern Africa, Parasites and Vectors. 11 (2018) 578. https://doi.org/10.1186/s13071-018-3163-3.

[28] The World Bank IBRD-IDA, Countries | Data, (2022). https://data.worldbank.org/country (accessed November 17, 2022).

[29] L. Phillips, Successful meat production on Réunion Island, Farmer’s Wkly. (2014). https://www.farmersweekly.co.za/animals/cattle/successful-meat-production-on-reunion-island/ (accessed September 27, 2022).

[30] H.C. Metz, Comoros: A Country Study, Washingt. GPO Libr. Congr. (1994). http://countrystudies.us/comoros/ (accessed September 27, 2022).

[31] A.A. Elmi, Livestock production in Somalia with special emphasis on camels, Nomad. People. 29 (1991) 87–103. https://www.jstor.org/stable/43123342 (accessed November 17, 2022).

[32] EAFF 2022, East Africa Farmers Federation, (n.d.). https://www.eaffu.org/djibouti/ (accessed September 23, 2022).

[33] Eritrea Ministry of Information, Livestock Population and Types, (2012). https://shabait.com/2012/12/24/livestock-population-and-types-of-eritrea/ (accessed September 27, 2022).

[34] MrMarc Naiken; Ms.Laura Ah Time, Seychelles Census of Agriculture, (2011). https://wedocs.unep.org/xmlui/handle/20.500.11822/9520 (accessed November 17, 2022).

[35] K.E. Mwape, S. Gabriël, The Parasitological, Immunological, and Molecular Diagnosis of Human Taeniasis with Special Emphasis on Taenia solium Taeniasis, Curr. Trop. Med. Reports. 1 (2014) 173–180. https://doi.org/10.1007/s40475-014-0028-5.

[36] H.H. Garcia, O.H. Del Brutto, Neurocysticercosis: Updated concepts about an old disease, Lancet Neurol. 4 (2005) 653–661. https://doi.org/10.1016/S1474-4422(05)70194-0.

[37] N.A. Mafojane, C.C. Appleton, R.C. Krecek, L.M. Michael, A.L. Willingham, The current status of neurocysticercosis in Eastern and Southern Africa, in: Acta Trop., Elsevier, 2003: pp. 25–33. https://doi.org/10.1016/S0001-706X(03)00052-4.

[38] H.J. Heinz, G.M. Macnab, Cysticercosis in the Bantu of southern Africa., S. Afr. J. Med. Sci. 30 (1965) 19–31. https://pubmed.ncbi.nlm.nih.gov/5854976/ (accessed May 15, 2022).

[39] I.K. Phiri, P. Dorny, S. Gabriel, A.L. Willingham, N. Speybroeck, J. Vercruysse, The prevalence of porcine cysticercosis in Eastern and Southern provinces of Zambia, Vet. Parasitol. 108 (2002) 31–39. https://doi.org/10.1016/S0304-4017(02)00165-6.

[40] F. Jansen, P. Dorny, S. Gabriël, V. Dermauw, M.V. Johansen, C. Trevisan, The survival and dispersal of Taenia eggs in the environment: what are the implications for transmission? A systematic review, Parasites and Vectors. 14 (2021) 1–16. https://doi.org/10.1186/s13071-021-04589-6.

[41] K.E. Mwape, J. Blocher, J. Wiefek, K. Schmidt, P. Dorny, N. Praet, C. Chiluba, H. Schmidt, I.K. Phiri, A.S. Winkler, S. Gabriël, Prevalence of neurocysticercosis in people with epilepsy in the Eastern province of Zambia, PLoS Negl. Trop. Dis. 9 (2015) e0003972. https://doi.org/10.1371/journal.pntd.0003972.

[42] A. Winkler, Sylvia, Epilepsy and Neurocysticercosis in Sub-Saharan Africa, Nov. Asp. Cysticercosis Neurocysticercosis. (2013). https://doi.org/10.5772/53289.

[43] A. Millogo, A. Kongnyu Njamnshi, M. Kabwa-PierreLuabeya, Neurocysticercosis and epilepsy in sub-Saharan Africa, Brain Res. Bull. 145 (2019) 30–38. https://doi.org/10.1016/j.brainresbull.2018.08.011.

[44] M. Laranjo-González, B. Devleesschauwer, C. Trevisan, A. Allepuz, S. Sotiraki, A. Abraham, M.B. Afonso, J. Blocher, L. Cardoso, J.M. Correia Da Costa, P. Dorny, S. Gabriël, J. Gomes, M.Á. Gómez-Morales, P. Jokelainen, M. Kaminski, B. Krt, P. Magnussen, L.J. Robertson, V. Schmidt, E. Schmutzhard, G.S.A. Smit, B. Šoba, C.R. Stensvold, J. Starič, K. Troell, A.V. Rataj, M. Vieira-Pinto, M. Vilhena, N.A. Wardrop, A.S. Winkler, V. Dermauw, Epidemiology of taeniosis/cysticercosis in Europe, a systematic review: Western Europe, Parasites and Vectors. 10 (2017) 1–14. https://doi.org/10.1186/s13071-017-2280-8.

[45] R. Baskind, G.L. Birbeck, Epilepsy-associated stigma in sub-Saharan Africa: The social landscape of a disease, Epilepsy Behav. 7 (2005) 68–73. https://doi.org/10.1016/J.YEBEH.2005.04.009.

[46] M. Atadzhanov, A. Haworth, E.N. Chomba, E.K. Mbewe, G.L. Birbeck, Epilepsy- associated stigma in Zambia: what factors predict greater felt stigma in a highly stigmatized population?, Epilepsy Behav. 19 (2010) 414–8. https://doi.org/10.1016/j.yebeh.2010.08.017.

[47] M.T. Medina, R.L. Aguilar-Estrada, A. Alvarez, R.M. Durõn, L. Martínez, S. Dubõn, A.L. Estrada, C. Zúniga, D. Cartagena, A. Thompson, E. Ramirez, L. Banegas, J.R. Osorio, A. V. Delgado-Escueta, J.S. Collins, K.R. Holden, Reduction in rate of epilepsy from neurocysticercosis by community interventions: The Salamá, Honduras study, Epilepsia. 52 (2011) 1177–1185. https://doi.org/10.1111/j.1528-1167.2010.02945.x.

[48] J.S. McCarthy, S. Lustigman, G.J. Yang, R.M. Barakat, H.H. García, B. Sripa, A.L. Willingham, R.K. Prichard, M.G. Basáñez, A research agenda for helminth diseases of humans: Diagnostics for control and elimination programmes, PLoS Negl. Trop. Dis. 6 (2012). https://doi.org/10.1371/journal.pntd.0001601.

[49] S. Thys, K.E. Mwape, P. Lefèvre, P. Dorny, T. Marcotty, A.M. Phiri, I.K. Phiri, S. Gabriël, Why Latrines Are Not Used: Communities’ Perceptions and Practices Regarding Latrines in a Taenia solium Endemic Rural Area in Eastern Zambia, PLoS Negl. Trop. Dis. 9 (2015) e0003570. https://doi.org/10.1371/journal.pntd.0003570.

[50] F.R. Rabenjamina, J. Ranaivoravo, V. Raholimina, A. Ramialiharisoa, A case report of bronchial cysticercosis, Arch. Inst. Pasteur Madagascar. 66 (2000) 43–45. https://europepmc.org/article/med/12463034 (accessed October 23, 2021).

[51] H.N. Rakoto-Ratsimba, S.S.E.N. Rabesalama, H.J.C. Razafimahandry, A. Ranaivozanany, Case report of solitary breast cysticercosis in Madagascar, Med. Trop. 67 (2007) 179–180. https://pubmed.ncbi.nlm.nih.gov/17691439/ (accessed October 18, 2021).

[52] T. Heller, C. Wallrauch, D. Kaminstein, S. Phiri, Case report: Cysticercosis: sonographic diagnosis of a treatable cause of epilepsy and skin nodules, Am. J. Trop. Med. Hyg. 97 (2017) 1827–1829. https://doi.org/10.4269/ajtmh.17-0257.

[53] S.J. Uledi, A rare gigantic solitary cysticercosis pseudotumour of the neck, J. Surg. Case Reports. 2010 (2010) 5–5. https://doi.org/10.1093/jscr/2010.9.5.

[54] S. Agnihotri, O.P. Talwar, S. Pudasaini, R. Baral, Cysticercosis of breast - A case report, Polish J. Pathol. 57 (2006) 53–54. https://doi.org/10.3348/jkrs.1995.32.5.835.

[55] J. Tuan, L. Kailani, P. Ngabitsinze, S. Umuganwa, F. Munyaneza, E. Musoni, A.L. Canales, M. Nkeshimana, Disseminated cysticercosis in rwanda—case report of a patient presenting with difficulty with walking and skin nodules, Rwanda Med. J. 77 (2020) 1–4. https://www.google.com/search?q=Disseminated+cysticercosis+in+rwanda—case+report+of+a+patient+presenting+with+difficulty+with+walking+and+skin+nodules&oq=Disseminated+cysticercosis+in+rwanda—case+report+of+a+patient+presenting+with+difficulty+with+walking (accessed July 5, 2022).

[56] M.B. Thomas, K.M. Thomas, A.A. Awotedu, E. Blanco-Blanco, M. Anwary, Cardiocysticercosis, SAMJ South African Med. J. 97 (2007) 504–506. https://go.gale.com/ps/i.do?p=AONE&sw=w&issn=02569574&v=2.1&it=r&id=GALE%7CA167881603&sid=googleScholar&linkaccess=fulltext (accessed October 24, 2021).

[57] E. Agaba, D. Modi, O. Gunduz, Z. Modi, Subcutaneous nodules of cysticercosis as a sign of asymptomatic neurocysticercosis in an HIV positive patient, Rev. Soc. Bras. Med. Trop. 51 (2018) 861–863. https://doi.org/10.1590/0037-8682-0178-2018.

[58] Z. Nowalaza, M. Zampoli, K. Pillay, S. Singh, H.J. Zar, Pulmonary cysticercosis in an urban South African child, Pediatr. Pulmonol. 56 (2021) 4060–4062. https://doi.org/10.1002/ppul.25682.

[59] D.W. McCormick, J.M. Bacha, N.K. El-Mallawany, C.L. Kovarik, J.S. Slone, L.R. Campbell, Disseminated cysticercosis and Kaposi sarcoma in a child with HIV/AIDS: A case report, BMC Infect. Dis. 20 (2020) 1–5. https://doi.org/10.1186/s12879-020-05039-x.

[60] F.R. Lyimo, A.M. Jusabani, H. Makungu, M. Mtolera, S. Surani, Thinking Outside Malaria: A Rare Case of Disseminated Cysticercosis With Cardiopulmonary Involvement From Urban Tanzania, Cureus. 13 (2021). https://doi.org/10.7759/cureus.12851.

[61] P. Rajaonarison, S. Ralamboson, C. Andriamamonjy, R. Ramanampamonjy, R. Ce, F. Razafindratrimo, R. Villeneuve, A. Andriantsimahavandy, Diagnostic de la neurocysticercose : à propos d ’ un cas, Arch. Inst. Pasteur Madagascar. 67 (2001) 53–56. https://www.researchgate.net/publication/237675160_Diagnostic_de_la_neurocysticercose_a_propos_d’un_cas (accessed October 23, 2021).

[62] R.A. Rakotoarivelo, R.L. Andrianasolo, D. Ranoharison, R.O.S. Rakoto, F.A. Sendrasoa, M.J. de Dieu Randria, Neurocysticercosis as an important differential of paradoxical response during antituberculosis therapy in HIV-negative patient, Asian Pacific J. Trop. Dis. 1 (2011) 333–334. https://doi.org/10.1016/S2222-1808(11)60077-7.

[63] N. Kalata, J. Ellis, L. Benjamin, S. Kampondeni, P. Chiodini, T. Harrison, D.G. Lalloo, R.S. Heyderman, Neurological deterioration in a patient with HIV-associated cryptococcal meningitis initially improving on antifungal treatment: a case report of coincidental racemose neurocysticercosis, BMC Infect. Dis. 21 (2021) 1–5. https://doi.org/10.1186/s12879-021-06425-9.

[64] V. Saldanha, G. Saldanha, R.P. Reys, C.A. Benson, E.V. Noormahomed, Neurocysticercosis in Child Bearing Women: An Overlooked Condition in Mozambique and a Potentially Missed Diagnosis in Women Presenting with Eclampsia., EC Microbiol. 14 (2018) 736–740. http://www.ncbi.nlm.nih.gov/pubmed/31681909%0Ahttp://www.pubmedcentral.nih.gov/articlerender.fcgi?artid=PMC6824723 (accessed July 5, 2022).

[65] A.M. Vitali, R. Jithoo, Multiple forms of neurocysticercosis in a single patient, Br. J. Neurosurg. 18 (2004) 650–651. https://doi.org/10.1080/02688690400022847.

[66] T. Motsepe, D. Ackerman, Spinal and vertebral neurocysticercosis in an HIV-positive female patient, South. African J. Epidemiol. Infect. 27 (2012) 133–136. https://doi.org/10.1080/10158782.2012.11441499.

[67] K. Jivan, A. Mochan, G. Modi, Intraventricular neurocysticercosis causing acute unilateral hydrocephalus, African J. Psychiatry (South Africa). 13 (2010) 315–317. https://www.researchgate.net/publication/47460261_Intraventricular_neurocysticercosis_causing_acute_unilateral_hydrocephalus (accessed October 22, 2021).

[68] H.F. Sibat, Neurocysticercosis, Epilepsy, COVID-19 and a Novel Hypothesis: Cases Series and Systematic Review, Clin. Schizophr. Relat. Psychoses. 15 (2021). https://doi.org/10.3371/CSRP.FH.121421.

[69] B. Ndlovu, Combined surgical and medical treatment of racemose neurocysticercosis in Swaziland, Neuroradiology. 60 (2018) 109–110. https://www.embase.com/search/results?subaction=viewrecord&id=L621458617&from=export.

[70] F. Mukonde, Abstracts, Epilepsia. 55 (2014) 4–246. https://doi.org/10.1111/epi.12675.

[71] J. Manske, C. McVey, H. Chengazi, O. Siddiqi, D. Saylor, Intramedullary spinal neurocysticercosis (P5.9-031), Neurology. 92 (2019).

[72] A. Musara, N. Soko, S. Shamu, Suprasellar cysticercosis cyst with optic nerve compression masquerading as an arachnoid cyst, Middle East Afr. J. Ophthalmol. 26 (2019) 114–116. https://doi.org/10.4103/meajo.MEAJO_142_18.

[73] G. Nsengiyumva, M. Druet-Cabanac, B. Ramanankandrasana, B. Bouteille, L. Nsizabira, P.M. Preux, Cysticercosis as a major risk factor for epilepsy in Burundi, East Africa, Epilepsia. 44 (2003) 950–955. https://doi.org/10.1046/j.1528-1157.2003.55302.x.

[74] P. Leutscher, A. Andriantsimahavandy, Cysticercosis in Peace Corps Volunteers in Madagascar, N. Engl. J. Med. 350 (2004) 311–312. https://doi.org/10.1056/nejm200401153500325.

[75] J.. Carod, J. Razafimahefa, M. Randrianarison, R.. Ramahefarisoa, M. Rakotondrazaka, M. Andriantseheno, The burden of neurocysticercosis: A multi-purpose survey in Madagascar, Clin. Microbiol. Infect. 18 (2012) 52–53. http://ovidsp.ovid.com/ovidweb.cgi?T=JS&PAGE=reference&D=emed10&NEWS=N&AN=70822228 (accessed July 8, 2022).

[76] J.. Carod, M. Rakotondrazaka, R.. Ramahefarisoa, D. Menard, J.. Soares, P. Dorny, P. Wilkins, Updated epidemiology of cysticercosis in Madagascar, Trop. Med. Int. Heal. 16 (2011) 270. http://ovidsp.ovid.com/ovidweb.cgi?T=JS&PAGE=reference&D=emed10&NEWS=N&AN=70589687 (accessed July 9, 2022).

[77] M.L. Rakotomahefa Narison, A.B.A. Ratsimbazafy, H. Randrianjafinimpanana, N. Randrianaivo, T.N. Andriatahina, H.S. Raobijaona, Aspect épidémio-clinique de la neurocysticercose dans le service de pédiatrie au CHU de Tamatave- Madagascar, Arch. Pediatr. 21 (2014) 810. https://doi.org/10.1016/S0929-693X(14)72070-6.

[78] N.J. Zafindraibe, J. Ralalarinivo, A.I. Rakotoniaina, M.N. Maeder, M.R. Andrianarivelo, B. Contamin, A. Michault, A. Rasamindrakotroka, Séroprévalence de la cysticercose et facteurs de risque associés chez un groupe de patients vus au centre hospitalier régional de référence d’antsirabe, madagascar, Pan Afr. Med. J. 28 (2017). https://doi.org/10.11604/pamj.2017.28.260.10463.

[79] J.F. Carod, P. Dorny, Cysticercosis in Madagascar, J. Infect. Dev. Ctries. 14 (2020) 1031–1042. https://doi.org/10.3855/JIDC.13450.

[80] Y.A. Assane, C. Trevisan, C.M. Schutte, E.V. Noormahomed, M.V. Johansen, P. Magnussen, Neurocysticercosis in a rural population with extensive pig production in Angónia district, Tete Province, Mozambique, Acta Trop. 165 (2017) 155–160. https://doi.org/10.1016/j.actatropica.2015.10.018.

[81] E.V. Noormahomed, N. Nhacupe, C. Mascaró-Lazcano, M.N. Mauaie, T. Buene, C.A. Funzamo, C.A. Benson, A Cross-sectional Serological Study of Cysticercosis, Schistosomiasis, Toxocariasis and Echinococcosis in HIV-1 Infected People in Beira, Mozambique, PLoS Negl. Trop. Dis. 8 (2014) e3121. https://doi.org/10.1371/journal.pntd.0003121.

[82] R. Rottbeck, J.F. Nshimiyimana, P. Tugirimana, U.E. Düll, J. Sattler, J.C. Hategekimana, J. Hitayezu, I. Bruckmaier, M. Borchert, J.B. Gahutu, S. Dieckmann, G. Harms, F.P. Mockenhaupt, R. Ignatius, High Prevalence of Cysticercosis in People with Epilepsy in Southern Rwanda, PLoS Negl. Trop. Dis. 7 (2013). https://doi.org/10.1371/journal.pntd.0002558.

[83] H. Foyaca-Sibat, L.D. Cowan, H. Carabin, I. Targonska, M.A. Anwary, G. Serrano- Ocaña, R.C. Krecek, A.L. Willingham, Accuracy of serological testing for the diagnosis of prevalent neurocysticercosis in outpatients with epilepsy, Eastern Cape Province, South Africa, PLoS Negl. Trop. Dis. 3 (2009) e562. https://doi.org/10.1371/journal.pntd.0000562.

[84] A.S. Winkler, J. Blocher, H. Auer, T. Gotwald, W. Matuja, E. Schmutzhard, Epilepsy and neurocysticercosis in rural Tanzania - An imaging study, Epilepsia. 50 (2009) 987–993. https://doi.org/10.1111/j.1528-1167.2008.01867.x.

[85] E. Hunter, K. Burton, A. Iqbal, D. Birchall, M. Jackson, J. Rogathe, A. Jusabani, W. Gray, E. Aris, G. Kamuyu, P.P. Wilkins, C.R. Newton, R. Walker, Cysticercosis and epilepsy in rural Tanzania: A community-based case-control and imaging study, Trop. Med. Int. Heal. 20 (2015) 1171–1179. https://doi.org/10.1111/tmi.12529.

[86] V. Schmidt, C. Kositz, K.H. Herbinger, H. Carabin, B. Ngowi, E. Naman, P.P. Wilkins, J. Noh, W. Matuja, A.S. Winkler, Association between Taenia solium infection and HIV/AIDS in northern Tanzania: A matched cross sectional-study, Infect. Dis. Poverty. 5 (2016). https://doi.org/10.1186/s40249-016-0209-7.

[87] V. Schmidt, M.C. O’Hara, B. Ngowi, K.H. Herbinger, J. Noh, P.P. Wilkins, V. Richter, C. Kositz, W. Matuja, A.S. Winkler, Taenia solium cysticercosis and taeniasis in urban settings: Epidemiological evidence from a health-center based study among people with epilepsy in Dar es Salaam, Tanzania, PLoS Negl. Trop. Dis. 13 (2019) e0007751. https://doi.org/10.1371/journal.pntd.0007751.

[88] J.O. Mageto, C.I. Muleke, C.R. Newton, G. Samuel, Multiple parasitic infections as risk factors of active convulsive epilepsy., Sch. J. Appl. Med. Sci. 3 (2015) 178–184. http://saspublisher.com/wp-content/uploads/2015/01/SJAMS-31C178-184.pdf.

[89] B.J. Mwang’Onde, M.J. Chacha, G. Nkwengulila, The status and health burden of neurocysticercosis in Mbulu district, northern Tanzania, BMC Res. Notes. 11 (2018) 1–5. https://doi.org/10.1186/s13104-018-3999-9.

[90] G. Kamuyu, C. Bottomley, J. Mageto, B. Lowe, P.P. Wilkins, J.C. Noh, T.B. Nutman, A.K. Ngugi, R. Odhiambo, R.G. Wagner, A. Kakooza-Mwesige, S. Owusu-Agyei, K. Ae- Ngibise, H. Masanja, F.H.A. Osier, P. Odermatt, C.R. Newton, Exposure to Multiple Parasites Is Associated with the Prevalence of Active Convulsive Epilepsy in Sub-Saharan Africa, PLoS Negl. Trop. Dis. 8 (2014) e2908. https://doi.org/10.1371/journal.pntd.0002908.

[91] K.E. Mwape, I.K. Phiri, N. Praet, N. Speybroeck, J.B. Muma, P. Dorny, S. Gabriël, The Incidence of Human Cysticercosis in a Rural Community of Eastern Zambia, PLoS Negl. Trop. Dis. 7 (2013) e2142. https://doi.org/10.1371/journal.pntd.0002142.

[92] J.J.J. Kumwenda, G. Mateyu, S. Kampondeni, A.P. Van Dam, L. Van Lieshout, E.E.E. Zijlstra, A. van Dam, L. van Lieshout, E.E.E. Zijlstra, Differential diagnosis of stroke in a setting of high HIV prevalence in Blantyre, Malawi, Stroke. 17 (2005) 960–964. https://doi.org/10.1161/01.STR.0000162585.97216.ef.

[93] I.L. Segamwenge, N.P. Kioko, C. Mukulu, O. Jacob, W. Humphrey, J. Augustinus, Neurocysticercosis among patients with first time seizure in Northern Namibia, Pan Afr. Med. J. 24 (2016). https://doi.org/10.11604/pamj.2016.24.127.8908.

[94] S.H. Mabaso, D. Bhana-Nathoo, S. Lucas, An audit of CT brain findings in adults with new-onset seizures in a resource restricted setting in South Africa, South African J. Radiol. 26 (2022) 7. https://doi.org/10.4102/SAJR.V26I1.2294.

[95] J.C. Sousa-Figueiredo, D. Gamboa, J.M. Pedro, C. Fançony, A.J. Langa, R.J.S. Magalhães, J.R. Stothard, S.V. Nery, Epidemiology of malaria, schistosomiasis, geohelminths, anemia and malnutrition in the context of a demographic surveillance system in northern Angola, PLoS One. 7 (2012) e33189. https://doi.org/10.1371/journal.pone.0033189.

[96] D. Oliveira, F.S. Ferreira, J. Atouguia, F. Fortes, A. Guerra, S. Centeno-Lima, Infection by intestinal parasites, stunting and anemia in school-aged children from southern Angola, PLoS One. 10 (2015) e0137327. https://doi.org/10.1371/journal.pone.0137327.

[97] S.T. Mariam, B. Roma, S. Sorsa, S. Worku, L. Erosie, Assessment of sanitary and hygienic status of catering establishments of Awassa Town, Ethiop. J. Heal. Dev. 14 (2000). https://doi.org/10.4314/ejhd.v14i1.9934.

[98] M. Awole, S. Gebre-Selassie, T. Kassa, G. Kibru, Prevalence of intestinal parasites in HIV-infected adult patients in Southwestern Ethiopia, Ethiop. J. Heal. Dev. 17 (2003). https://doi.org/10.4314/ejhd.v17i1.9783.

[99] Z. Mekonnen, S. Suleman, A. Biruksew, T. Tefera, L. Chelkeba, Intestinal polyparasitism with special emphasis to soil-transmitted helminths among residents around Gilgel Gibe Dam, Southwest Ethiopia: a community based survey, BMC Public Health. 16 (2016) 1–7. https://doi.org/10.1186/s12889-016-3859-2.

[100] A. Alemu, Y. Shiferaw, G. Getnet, A. Yalew, Z. Addis, Opportunistic and other intestinal parasites among HIV/AIDS patients attending Gambi higher clinic in Bahir Dar city, North West Ethiopia, Asian Pac. J. Trop. Med. 4 (2011) 661–665. https://doi.org/10.1016/S1995-7645(11)60168-5.

[101] T. Wegayehu, T. Tsalla, B. Seifu, T. Teklu, Prevalence of intestinal parasitic infections among highland and lowland dwellers in Gamo area, South Ethiopia, BMC Public Health. 13 (2013) 1–7. https://doi.org/10.1186/1471-2458-13-151.

[102] M. Alamir, W. Awoke, A. Feleke, Intestinal parasites infection and associated factors among school children in Dagi primary school, Amhara National Regional State, Ethiopia, Health (Irvine. Calif). 05 (2013) 1697–1701. https://doi.org/10.4236/health.2013.510228.

[103] S. Fekadu, K. Taye, W. Teshome, S. Asnake, Prevalence of parasitic infections in HIV- positive patients in southern Ethiopia: A cross-sectional study, J. Infect. Dev. Ctries. 7 (2013) 868–872. https://doi.org/10.3855/jidc.2906.

[104] B. Abera, G. Alem, M. Yimer, Z. Herrador, Epidemiology of soil-transmitted helminths, Schistosoma mansoni, and haematocrit values among schoolchildren in Ethiopia, J. Infect. Dev. Ctries. 7 (2013) 253–260. https://doi.org/10.3855/jidc.2539.

[105] H. Mamo, Intestinal parasitic infections among prison inmates and tobacco farm workers in Shewa Robit, north-central Ethiopia, PLoS One. 9 (2014) e99559. https://doi.org/10.1371/journal.pone.0099559.

[106] L. Worku, D. Damte, M. Endris, H. Tesfa, M. Aemero, Schistosoma mansoni infection and associated determinant factors among school children in Sanja Town, northwest Ethiopia, J. Parasitol. Res. 2014 (2014). https://doi.org/10.1155/2014/792536.

[107] M. Alemayehu, W. Birhan, Y. Belyhun, M. Sahle, B. Tessema, Prevalence of smear positive tuberculosis, intestinal parasites and their co-infection among tuberculosis suspects in Gondar University Hospital and Gondar Poly Clinic, North West Ethiopia, J. Microb. Biochem. Technol. 6 (2014) 179–184. https://doi.org/10.4172/1948-5948.1000140.

[108] M. Desta, D. Asrat, W. Yimtubezinash, N. Demiss, Prevalence of intestinal parasites and Salmonella and Shigella among food handlers at food service establishments in the main campus and Health Sciences College of Hawassa University, Hawassa, Ethiopia, Ethiop. J. Heal. Dev. 28 (2014) 29–34. http://www.etpha.org/publications/the-ethiopian-journal-of-health-development.html%0Ahttp://ovidsp.ovid.com/ovidweb.cgi?T=JS&CSC=Y&NEWS=N&PAGE=fulltext&D=cagh&AN=20153041959%0Ahttp://wa4py6yj8t.search.serialssolutions.com/?url_ver=Z39.88-2004&rft_val_fmt= (accessed October 18, 2021).

[109] G. Beyene, H. Tasew, Prevalence of intestinal parasite, Shigella and Salmonella species among diarrheal children in Jimma health center, Jimma southwest Ethiopia: A cross sectional study, Ann. Clin. Microbiol. Antimicrob. 13 (2014) 10. https://doi.org/10.1186/1476-0711-13-10.

[110] A. Abossie, M. Seid, Assessment of the prevalence of intestinal parasitosis and associated risk factors among primary school children in Chencha town, Southern Ethiopia, BMC Public Health. 14 (2014). https://doi.org/10.1186/1471-2458-14-166.

[111] A. Gebreyesus, K. Adane, L. Negash, T. Asmelash, S. Belay, M. Alemu, M. Saravanan, Prevalence of Salmonella typhi and intestinal parasites among food handlers in Mekelle University student cafeteria, Mekelle, Ethiopia, Food Control. 44 (2014) 45–48. https://doi.org/10.1016/j.foodcont.2014.03.040.

[112] J.M. Ramos, N. Rodríguez-Valero, G. Tisiano, H. Fano, T. Yohannes, A. Gosa, E. Fruttero, F. Reyes, M. Górgolas, Different profile of intestinal protozoa and helminthic infections among patients with diarrhoea according to age attending a rural hospital in southern Ethiopia, Trop. Biomed. 31 (2014) 392–397. https://pubmed.ncbi.nlm.nih.gov/25134911/ (accessed October 27, 2021).

[113] M.B. Shiferaw, A.D. Mengistu, Helminthiasis: Hookworm infection remains a public health problem in Dera District, South Gondar, Ethiopia, PLoS One. 10 (2015) e0144588. https://doi.org/10.1371/journal.pone.0144588.

[114] A. Bereket, T. Zewdneh, Prevalence of intestinal helminthiasis and associated risk factors among schoolchildren in Dawro Zone, Southern Ethiopia, J. Biol. Agric. Healthc. 5 (2015) 76–82. https://www.iiste.org/Journals/index.php/JBAH/article/view/23238 (accessed October 27, 2021).

[115] G. Tigist, E. Fantahun, E. Hailu, H. Kibe, O. Fesseha, W. Godana, W.G. Tsadik, Prevalence of Intestinal Parasitosis Among HIV / AIDS Patients Attending ART Clinic of Arbaminch Hospital., Researchgate.Net. (2015) 1–6. https://www.researchgate.net/profile/Wanzahun-Godana/publication/283351146_PREVALENCE_OF_INTESTINAL_PARASITOSIS_AMONG_HIVAIDS_PATIENTS_ATTENDING_ART_CLINIC_OF_ARBAMINCH_HOSPITAL/links/5636572908aeb786b703f0ae/PREVALENCE-OF-INTESTINAL-PARASITOSIS-AMONG-HIV (accessed October 22, 2021).

[116] M. Mama, G. Alemu, Prevalence and factors associated with intestinal parasitic infections among food handlers of Southern Ethiopia: Cross sectional study Infectious Disease epidemiology, BMC Public Health. 16 (2016) 1–7. https://doi.org/10.1186/s12889-016-2790-x.

[117] A. Alemu, Y. Tegegne, D. Damte, M. Melku, Schistosoma mansoni and soil-transmitted helminths among preschool-aged children in Chuahit, Dembia district, Northwest Ethiopia: Prevalence, intensity of infection and associated risk factors, BMC Public Health. 16 (2016). https://doi.org/10.1186/s12889-016-2864-9.

[118] A. Arota Amado, F. Wadilo Wada, F. Solomon Bisetegn, Y. Abreham Leka, Non- opportunistic intestinal parasitic infections among HIV-infected individuals at Wolaita Sodo Hospital, South Ethiopia, J. Coast. Life Med. 4 (2016) 353–357. https://doi.org/10.12980/jclm.4.2016j6-57.

[119] B. Alemayehu, Z. Tomass, F. Wadilo, D. Leja, S. Liang, B. Erko, Epidemiology of intestinal helminthiasis among school children with emphasis on Schistosoma mansoni infection in Wolaita zone, Southern Ethiopia, BMC Public Health. 17 (2017). https://doi.org/10.1186/s12889-017-4499-x.

[120] H. Girma, G. Beyene, Z. Mekonnen, Prevalence of intestinal parasites among food handlers at cafeteria of Jimma University Specialized Hospital, Southwest Ethiopia, Asian Pacific J. Trop. Dis. 7 (2017) 467–471. https://doi.org/10.12980/apjtd.7.2017D7-20.

[121] M. Alemu, B. Kinfe, D. Tadesse, W. Mulu, T. Hailu, E. Yizengaw, Intestinal parasitosis and anaemia among patients in a Health Center, North Ethiopia, BMC Res. Notes. 10 (2017) 1–7. https://doi.org/10.1186/s13104-017-2957-2.

[122] D.G. Feleke, S. Arega, M. Tekleweini, K. Kindie, A. Gedefie, Schistosoma mansoni and other helminthes infections at Haike primary school children, North-East, Ethiopia: A cross-sectional study, BMC Res. Notes. 10 (2017) 1–6. https://doi.org/10.1186/s13104-017-2942-9.

[123] Y. Tegegne, T. Wondmagegn, L. Worku, A. Jejaw Zeleke, Prevalence of Intestinal Parasites and Associated Factors among Pulmonary Tuberculosis Suspected Patients Attending University of Gondar Hospital, Gondar, Northwest Ethiopia, J. Parasitol. Res. 2018 (2018). https://doi.org/10.1155/2018/9372145.

[124] G. Alemu, M. Mama, Intestinal helminth co-infection and associated factors among tuberculosis patients in Arba Minch, Ethiopia, BMC Infect. Dis. 17 (2017) 1–9. https://doi.org/10.1186/s12879-017-2195-1.

[125] T. Asmare, W. Awoke, G. Alem, Prevalence of Intestinal Parasites and Associated Factors among AdultPre-ART and ART Patients in Goncha Siso Enesie Woreda, East Gojjam,Northwest Ethiopia, 2014, J. AIDS Clin. Res. 8 (2017). https://doi.org/10.4172/2155-6113.1000734.

[126] D. Teklemariam, M. Legesse, A. Degarege, S. Liang, B. Erko, Schistosoma mansoni and other intestinal parasitic infections in schoolchildren and vervet monkeys in Lake Ziway area, Ethiopia, BMC Res. Notes. 11 (2018) 1–6. https://doi.org/10.1186/s13104-018-3248-2.

[127] T. Zemene, M.B. Shiferaw, Prevalence of intestinal parasitic infections in children under the age of 5 years attending the Debre Birhan referral hospital, North Shoa, Ethiopia, BMC Res. Notes. 11 (2018) 1–6. https://doi.org/10.1186/s13104-018-3166-3.

[128] G. Alemu, Z. Aschalew, E. Zerihun, Burden of intestinal helminths and associated factors three years after initiation of mass drug administration in Arbaminch Zuria district, Southern Ethiopia, BMC Infect. Dis. 18 (2018). https://doi.org/10.1186/s12879-018-3330-3.

[129] M. Alemu, A. Anley, K. Tedla, Magnitude of Intestinal Parasitosis and Associated Factors in Rural School Children, Northwest Ethiopia, Ethiop. J. Health Sci. 29 (2019) 923–928. https://doi.org/10.4314/ejhs.v29i1.14.

[130] B. Sitotaw, H. Mekuriaw, D. Damtie, Prevalence of intestinal parasitic infections and associated risk factors among Jawi primary school children, Jawi town, north-west Ethiopia, BMC Infect. Dis. 19 (2019). https://doi.org/10.1186/s12879-019-3971-x.

[131] E. Kebede, A. Seid, S. Akele, Prevalence and associated risk factors of intestinal parasitic infections among asymptomatic food handlers in Wollo University student’s cafeteria, Northeastern Ethiopia, BMC Res. Notes. 12 (2019) 1–6. https://doi.org/10.1186/s13104-019-4182-7.

[132] T. Eyamo, M. Girma, T. Alemayehu, Z. Bedewi, Soil-Transmitted Helminths And Other Intestinal Parasites Among Schoolchildren In Southern Ethiopia, Res. Rep. Trop. Med. Volume 10 (2019) 137–143. https://doi.org/10.2147/rrtm.s210200.

[133] N. Mohamed, A. Muse, M. Wordofa, D. Abera, A. Mesfin, M. Wolde, K. Desta, A. Tsegaye, B. Taye, Increased prevalence of cestode infection associated with history of deworming among primary school children in Ethiopia, Am. J. Trop. Med. Hyg. 101 (2019) 641–649. https://doi.org/10.4269/ajtmh.19-0284.

[134] A.A. Obala, C.J. Simiyu, D.O. Odhiambo, V. Nanyu, P. Chege, R. Downing, E. Mwaliko, A.W. Mwangi, D. Menya, D. Chelagat, H.D.N. Nyamogoba, P.O. Ayuo, W.P. O’Meara, M. Twagirumukiza, D. Vandenbroek, B.B.O. Otsyula, J. De Maeseneer, Webuye health and demographic surveillance systems baseline survey of soil-transmitted helminths and intestinal protozoa among children up to five years, J. Trop. Med. 2013 (2013). https://doi.org/10.1155/2013/734562.

[135] B.N. Lowoko, B. Ngugi, A. Malai, J.K. Mutai, Factors associated with intestinal parasites among school going children in Lodwar Municipality, Turkana County, Kenya, East Afr. Med. J. 94 (2018) 649–663. https://doi.org/10.4314/eamj.v94i8.

[136] T.P.W. Jones, J.D. Hart, K. Kalua, R.L. Bailey, A prevalence survey of enteral parasites in preschool children in the Mangochi District of Malawi, BMC Infect. Dis. 19 (2019). https://doi.org/10.1186/s12879-019-4439-8.

[137] E. V. Noormahomed, J.G. Pividal, S. Azzouz, C. Mascaró, M. Delgado-Rodríguez, A. Osuna, Seroprevalence of anti-cysticercus antibodies among the children living in the urban environs of Maputo, Mozambique, Ann. Trop. Med. Parasitol. 97 (2003) 31–35. https://doi.org/10.1179/000349803125002742.

[138] G. Augusto, R. Nalá, V. Casmo, A. Sabonete, L. Mapaco, J. Monteiro, Geographic distribution and prevalence of schistosomiasis and soil-transmitted helminths among schoolchildren in mozambique, Am. J. Trop. Med. Hyg. 81 (2009) 799–803. https://doi.org/10.4269/ajtmh.2009.08-0344.

[139] A.R.P. Walker, L.A. Dini, B.F. Walker, J.A. Frean, Helminthiasis in African children in a relatively low risk region in south Africa: implications for treatment?, S Afr J Epidemiol Infect South. 15 (2000). https://www.cabdirect.org/cabdirect/abstract/20013078709 (accessed October 22, 2021).

[140] H. Sacolo-Gwebu, M. Chimbari, C. Kalinda, Prevalence and risk factors of schistosomiasis and soil-transmitted helminthiases among preschool aged children (1-5 years) in rural KwaZulu-Natal, South Africa: A cross-sectional study, Infect. Dis. Poverty. 8 (2019) 1–12. https://doi.org/10.1186/s40249-019-0561-5.

[141] U.C. Braae, P. Magnussen, W. Harrison, B. Ndawi, F. Lekule, M.V. Johansen, Effect of National Schistosomiasis Control Programme on Taenia solium taeniosis and porcine cysticercosis in rural communities of Tanzania, Parasite Epidemiol. Control. 1 (2016) 245–251. https://doi.org/10.1016/j.parepi.2016.08.004.

[142] E. Buzigi, Prevalence of Intestinal Parasites , and its Association with Severe Acute Malnutrition Related Diarrhoea, J. Biol. Agric. Healthc. 5 (2015). https://www.researchgate.net/publication/271763982_Prevalence_of_Intestinal_Parasites_and_its_Association_with_Severe_Acute_Malnutrition_Related_Diarrhoea (accessed October 18, 2021).

[143] S. Fuhrimann, M.S. Winkler, N.B. Kabatereine, E.M. Tukahebwa, A.A. Halage, E. Rutebemberwa, K. Medlicott, C. Schindler, J. Utzinger, G. Cissé, Risk of Intestinal Parasitic Infections in People with Different Exposures to Wastewater and Fecal Sludge in Kampala, Uganda: A Cross-Sectional Study, PLoS Negl. Trop. Dis. 10 (2016) e0004469. https://doi.org/10.1371/journal.pntd.0004469.

[144] P. Oboth, Y. Gavamukulya, B.J. Barugahare, Prevalence and clinical outcomes of Plasmodium falciparum and intestinal parasitic infections among children in Kiryandongo refugee camp, mid-Western Uganda: A cross sectional study, BMC Infect. Dis. 19 (2019) 1–8. https://doi.org/10.1186/s12879-019-3939-x.

[145] K. Modjarrad, I. Zulu, D.T. Redden, L. Njobvu, D.O. Freedman, S.H. Vermund, Prevalence and predictors of intestinal helminth infections among human immunodeficiency virus type 1-infected adults in an urban African setting, Am. J. Trop. Med. Hyg. 73 (2005) 777–782. https://doi.org/10.4269/ajtmh.2005.73.777.

[146] J. Siwila, I.G.K. Phiri, H.L. Enemark, M. Nchito, A. Olsen, Intestinal helminths and protozoa in children in pre-schools in Kafue district, Zambia, Trans. R. Soc. Trop. Med. Hyg. 104 (2010) 122–128. https://doi.org/10.1016/j.trstmh.2009.07.024.

