## Supplement File 1 for "The epidemiology of human *Taenia solium* infections: a systematic review of the distribution in Eastern and Southern Africa"

**Documentation on the literature search for: The epidemiology of human Taenia solium taeniosis /cysticercosis: a systematic review of the distribution in Eastern and Southern Africa**

The following databases were searched:

| **Database** | **Number of retrieved records** |
| --- | --- |
| MEDLINE (Ovid) | 389 |
| Embase (Ovid) | 573 |
| Global Health (Ovid): | 561 |
| Scopus (Elsevier) | 575 |
| African Index Medicus (via WHO Global Index Medicus) | 8 |
| OpenGrey | 8 |
| Number of records before deduplication: | 2114 |
| Number of duplicate records removed: | 1198 |
| Number of records after deduplication: | 916 |

All searches were performed by Hilde Stromme, Academic Librarian, University of Oslo Library of Medicine and Science on June 20th, 2022. Exeption: The search in OpenGrey was performed on June 18th 2020. OpenGrey was closed in early 2021.

**Ovid MEDLINE(R) ALL <1946 to June 17, 2022>**

Date searched: 20 June 2022

| 1 | Taenia solium/ or exp Cysticercosis/ | 6703 |
| --- | --- | --- |
| 2 | (cysticercus cellulosae or ((taenia or t) adj solium*) or ((pork or porcine or pig*) adj2 (tapeworm* or worm*)) or cysticerc* or cisticerc* or neurocysticerc* or neurocisticerc*).tw,kf. | 8739 |
| 3 | 1 or 2 | 9358 |
| 4 | exp "Africa, Southern"/ or exp "Africa, Eastern"/ | 146116 |
| 5 | ((South* adj3 Africa*) or (East* adj3 Africa*) or Angola or Botswana or Kalahari or Burundi or Urundi or Comoro* or Djibouti or Somaliland or Eritrea or Ethiopia or Kenya or Lesotho or Madagascar or Malagasy or Malawi or Mauritius or Mayotte or Mozambique or Mocambique or Namibia or Reunion or Rwanda or Ruanda or Seychelles or Somalia or Somaliland or South Africa or Sudan or Swaziland or Eswatini or Tanzania or Zanzibar or Uganda or Zambia or Zimbabwe).tw,kf,kw,in. | 298782 |
| 6 | 4 or 5 | 325123 |
| 7 | 3 and 6 | 550 |
| 8 | limit 7 to yr="2000 - 2022" | 389 |

**Embase Classic+Embase <1947 to 2022 June 17>**

Date searched: 20 June 2022

| 1 | Taenia solium/ or cysticercosis/ or neurocysticercosis/ | 9883 |
| --- | --- | --- |
| 2 | (cysticercus cellulosae or ((taenia or t) adj solium*) or ((pork or porcine or pig*) adj2 (tapeworm* or worm*)) or cysticerc* or cisticerc* or neurocysticerc* or neurocisticerc*).tw,kf,kw. | 10414 |
| 3 | 1 or 2 | 12197 |
| 4 | Angola/ or Botswana/ or Burundi/ or Comoros/ or Djibouti/ or Eritrea/ or Ethiopia/ or Kenya/ or Lesotho/ or Malawi/ or Mauritius/ or Mayotte/ or Mozambique/ or Namibia/ or Reunion/ or Rwanda/ or Seychelles/ or Somalia/ or Somaliland/ or South Africa/ or Sudan/ or South Sudan/ or Eswatini/ or Swaziland/ or Tanzania/ or Uganda/ or Zambia/ or Zimbabwe/ or exp East African/ or exp Southern African/ | 192498 |
| 5 | ((South* adj3 Africa*) or (East* adj3 Africa*) or Angola or Botswana or Kalahari or Burundi or Urundi or Comoro* or Djibouti or Somaliland or Eritrea or Ethiopia or Kenya or Lesotho or Madagascar or Malagasy or Malawi or Mauritius or Mayotte or Mozambique or Mocambique or Namibia or Reunion or Rwanda or Ruanda or Seychelles or Somalia or Somaliland or South Africa or Sudan or Swaziland or Eswatini or Tanzania or Zanzibar or Uganda or Zambia or Zimbabwe).tw,kf,kw,in. | 397450 |
| 6 | 4 or 5 | 422694 |
| 7 | 3 and 6 | 769 |
| 8 | limit 7 to yr="2000 - 2022" | 573 |

**Global Health <1973 to 2022 Week 24>**

Date searched: 20 June 2022

| 1 | Taenia solium/ or cysticercosis/ or neurocysticercosis/ | 6482 |
| --- | --- | --- |
| 2 | (cysticercus cellulosae or ((taenia or t) adj solium*) or ((pork or porcine or pig*) adj2 (tapeworm* or worm*)) or cysticerc* or cisticerc* or neurocysticerc* or neurocisticerc*).dt,ti,ab,hw. | 8123 |
| 3 | 1 or 2 | 8123 |
| 4 | exp East Africa/ or exp Southern Africa/ | 116946 |
| 5 | ((South* adj3 Africa*) or (East* adj3 Africa*) or Angola or Botswana or Kalahari or Burundi or Urundi or Comoro* or Djibouti or Somaliland or Eritrea or Ethiopia or Kenya or Lesotho or Madagascar or Malagasy or Malawi or Mauritius or Mayotte or Mozambique or Mocambique or Namibia or Reunion or Rwanda or Ruanda or Seychelles or Somalia or Somaliland or South Africa or Sudan or Swaziland or Eswatini or Tanzania or Zanzibar or Uganda or Zambia or Zimbabwe).dt,ti,ab,hw,in,cp. | 254338 |
| 6 | 4 or 5 | 254338 |
| 7 | 3 and 6 | 913 |
| 8 | limit 7 to yr="2000 - 2022" | 561 |

**Scopus (Elsevier)**

TITLE-ABS-KEY (("cysticercus cellulosae" OR ((taenia OR t) PRE/0 solium*) OR ((pork OR porcine OR pig*) W/1 tapeworm* OR worm*) OR cysticerc* OR cisticerc* OR neurocysticerc* OR neurocisticerc*)) AND ((TITLE-ABS-KEY (((south* W/2 africa*) OR (east* W/2 africa*) OR angola OR botswana OR kalahari OR burundi OR urundi OR comoro* OR djibouti OR somaliland OR eritrea OR ethiopia OR kenya OR lesotho OR madagascar OR malagasy OR malawi OR mauritius OR mayotte OR mozambique OR mocambique OR namibia OR reunion OR rwanda OR ruanda OR seychelles OR somalia OR somaliland OR sudan OR swaziland OR eswatini OR tanzania OR zanzibar OR uganda OR zambia OR zimbabwe))) OR (AFFILCOUNTRY (((south* W/2 africa*) OR (east* W/2 africa*) OR angola OR botswana OR kalahari OR burundi OR urundi OR comoro* OR djibouti OR somaliland OR eritrea OR ethiopia OR kenya OR lesotho OR madagascar OR malagasy OR malawi OR mauritius OR mayotte OR mozambique OR mocambique OR namibia OR reunion OR rwanda OR ruanda OR seychelles OR somalia OR somaliland OR sudan OR swaziland OR eswatini OR tanzania OR zanzibar OR uganda OR zambia OR zimbabwe)))) AND (LIMIT-TO (PUBYEAR , 2022) OR LIMIT-TO (PUBYEAR , 2021) OR LIMIT-TO (PUBYEAR , 2020) OR LIMIT-TO (PUBYEAR , 2019) OR LIMIT-TO (PUBYEAR , 2018) OR LIMIT-TO (PUBYEAR , 2017) OR LIMIT-TO (PUBYEAR , 2016) OR LIMIT-TO (PUBYEAR , 2015) OR LIMIT-TO (PUBYEAR , 2014) OR LIMIT-TO (PUBYEAR , 2013) OR LIMIT-TO (PUBYEAR , 2012) OR LIMIT-TO (PUBYEAR , 2011) OR LIMIT-TO (PUBYEAR , 2010) OR LIMIT-TO (PUBYEAR , 2009) OR LIMIT-TO (PUBYEAR , 2008) OR LIMIT-TO (PUBYEAR , 2007) OR LIMIT-TO (PUBYEAR , 2006) OR LIMIT-TO (PUBYEAR , 2005) OR LIMIT-TO (PUBYEAR , 2004) OR LIMIT-TO (PUBYEAR , 2003) OR LIMIT-TO (PUBYEAR , 2002) OR LIMIT-TO (PUBYEAR , 2001) OR LIMIT-TO (PUBYEAR , 2000))

575 hits

**African Index Medicus**

(mh:("taenia solium")) OR (mh:("neurocysticercosis")) OR (tw:(cysticerc* OR cisticerc* OR neurocysticerc* OR neurocisticerc* OR "cysticercus cellulosae" OR "taenia solium"))

Limited to publication years 2000-2022.

8 hits.

**OpenGrey**
Date searched: 18 June 2020
Number of hits: 8

(cysticercus cellulosae OR ((taenia OR t) NEAR/0 solium*) OR ((pork OR porcine OR pig*) NEAR/1 tapeworm* OR worm*) OR cysticerc* OR cisticerc* OR neurocysticerc* OR neurocisticerc*) AND ((South* NEAR/2 Africa*) OR (East* NEAR/2 Africa*) OR Angola OR Botswana OR Kalahari OR Burundi OR Urundi OR Comoro* OR Djibouti OR Somaliland OR Eritrea OR Ethiopia OR Kenya OR Lesotho OR Madagascar OR Malagasy OR Malawi OR Mauritius OR Mayotte OR Mozambique OR Mocambique OR Namibia OR Reunion OR Rwanda OR Ruanda OR Seychelles OR Somalia OR Somaliland OR South Africa OR Sudan OR Swaziland OR Eswatini OR Tanzania OR Zanzibar OR Uganda OR Zambia OR Zimbabwe)
