## Supplement File 2 for "The epidemiology of human *Taenia solium* infections: a systematic review of the distribution in Eastern and Southern Africa"

**Data collection forms**

**Table: Individual human (neuro)cysticercosis cases**

| **Country** | **Date of detection** | **Age** | **Gender** | **Sample** | **Detection method** | **Location of pathogen** | **Comment** | **Reference** |
| --- | --- | --- | --- | --- | --- | --- | --- | --- |

**Table: Aggregated human (neuro)cysticercosis cases**

| **Year** | **Country in which the study conducted** | **Number of cases (prevalence in %)** | **Number of people tested** | **Detection method** | | | **Comment** | **Reference** |
| --- | --- | --- | --- | --- | --- | --- | --- | --- |
|  |  |  |  | **Immunology** | **Imaging** | **Other** |  |  |

**Table: Aggregated human taeniosis cases**

| **year** | **Country in which the study conducted** | **Number of cases (Prevalence in %)** | **Number of people tested** | **Sample** | **Detection method** | **Taenia species reported as** | **Comment** | **Reference** |
| --- | --- | --- | --- | --- | --- | --- | --- | --- |
